# Genome-wide methylation profiling identify hypermethylated HOXL subclass genes as potential markers for esophageal squamous cell carcinoma detection

**DOI:** 10.1101/2022.05.11.22274930

**Authors:** Qiuning Yu, Namei Xia, Yanteng Zhao, Huifang Jin, Renyin Chen, Fanglei Ye, Liyinghui Chen, Ying Xie, Kangkang Wan, Jun Zhou, Dihan Zhou, Xianping Lv

## Abstract

**Background:** Numerous studies have revealed aberrant DNA methylation in esophageal squamous cell carcinoma (ESCC). However, they often focused on the partial genome, which resulted in an inadequate understanding of the shaped methylation features and the lack of available methylation markers for this disease.

**Methods:** The current study investigated the methylation profiles between ESCC and paired normal samples using whole-genome bisulfite sequencing (WGBS) data and obtained a group of differentially methylated CpGs (DMC), differentially methylated regions (DMR), and differentially methylated genes (DMG). The DMGs were then verified in independent datasets and Sanger sequencing in our custom samples. Finally, we attempted to evaluate the performance of these genes as methylation markers for the classification of ESCC and multiple cancer types.

**Results:** We obtained 438,558 DMCs, 15,462 DMRs, and 1,568 DMGs. The four significantly enriched gene families of DMGs were CD molecules, NKL subclass, HOXL subclass, and Zinc finger C2H2-type. The HOXL subclass homeobox genes were observed extensively hypermethylated in ESCC and nine other cancer types. The HOXL-score estimated by *HOXC10* and *HOXD1* methylation showed good ability in discriminating ESCC from normal samples, while the methylation of *GSX1* displayed potential utility for pan-cancer detection.

**Conclusions:** We observed widespread hypomethylation events in ESCC, and the hypermethylated HOXL subclass homeobox genes presented promising applications for the early detection of multiple cancer types.

## Background

Esophageal cancer (EC) is one of the top 10 fetal malignant tumors worldwide, with a five-year overall survival rate of less than 20% [1]. The incidence of EC in men is four to five times higher than in women, and it predominantly occurs in East Asia as well as Eastern and Southern Africa [2]. Esophageal adenocarcinoma (ESCA) and esophageal squamous carcinoma (ESCC) are two major histologic subtypes of EC, with ESCC being the most common type [3]. Risk factors for developing EC are complex and vary among different histologic subtypes. ESCA is prevalent in Caucasian populations, and risk factors include obesity, gastroesophageal reflux disease, and Barrett’s esophagus. In contrast, ESCC is the predominant type of EC in East Asia and sub-Saharan Africa [4], while its main risk factors are papillomavirus infection, smoking, alcohol consumption, and hot foods [5]. Currently, EC is the fourth leading cause of cancer-related deaths in China, and ESCC is the most frequently diagnosed type (accounting for more than 90% of all EC cases), which is different from that in Western countries [6]. The morbidity and mortality of ESCC in China increase with age. The disease risk rapidly escalates after the age of 40, and the mortality rises after the age of 50 in the population [7].

Various staging strategies have been proposed to better guide the clinical management of esophageal cancer, and the TNM staging criteria (8th edition) jointly developed by the American Joint Commission on Cancer and the Union for International Cancer Control in 2017 is one of the widely used references [8]. In addition, NCCN has also released the clinical practice guidelines for esophageal cancer [9]. Accurately staging ESCC is critical for the clinical management of this disease. According to the eighth edition TNM categories, patients with lesions < 2 cm, tumors limited in the mucosal lamina propria or muscularis mucosae, moderate to high differentiation, and low risk of lymph node metastasis, local recurrence, or distant metastasis are classified as early stage. The high mortality and low 5-year survival rate of ESCC are mainly attributed to its advanced stage at diagnosis. The 5-year survival rate of early-stage ESCC improved from less than 20% to 80-90% after surgical or endoscopic resection [10]. Therefore, early detection can help reduce the incidence of ESCC and prolongs patient prognostic survival time. Traditionally, endoscopy has been the first choice for ESCC screening and can detect intraepithelial neoplasia, such as dysplasia and local non-invasive carcinoma, in asymptomatic patients, which is recognized as the major precancerous lesions [11]. A long-term follow-up showed that appropriate treatment of esophageal squamous epithelial dysplasia and early-stage ESCC after endoscopic screening reduced the disease mortality at the average-risk population in China [12]. In most developing countries, however, extensive endoscopic screening is not feasible given the cost-effectiveness. In contrast, inexpensive non-endoscopic esophageal sampling methods were proposed for ESCC screening. These sampling techniques combined with cytological examination have displayed some advantages, although the results of several studies are not satisfactory [13]. The potential utility of molecular diagnostic markers has been demonstrated for early detection of EC, including cell-free miRNAs and genomic abnormally methylated DNA [14,15]. Although the combination of DNA methylation and esophageal sampling techniques has presented a high accuracy in discriminating ESCC from normal controls [16,17], available methylation markers are still inadequate, and minimally invasive detection techniques based on blood methylation markers are urgently needed to be developed.

Several abnormal methylated genes on ESCC have been reported so far, and they are grouped to DNA damage repair, cell cycle regulation, cell adhesion, proliferation, and other biological categories [18]. These identified hypermethylated genes include *MGMT, MLH1* (DNA damage repair), *CDKN2A, CHFR* and *CDKN2B* (cell cycle regulation), *APC* and *SOX17* (Wnt signaling pathway), *RUNX3* and *DACH1* (transforming growth factor -β), and *CDH1, TFF1, TFPI2* (other biological functions). Previous studies suggested that some hypermethylated genes occurred in early-stage ESCC, including the well-known genes *MGMT* [19], *CDKN2A* [20], *MLH1* [21], and *CDH1* [22], while some are hypermethylated in late-stage such as *CHFR* [23], and others, such as hypermethylated *APC* are not associated with ESCC stage [24]. Although these genes are reported to be significantly hypermethylated on ESCC, their hypermethylation frequencies are not satisfied (from 30% to 60%) [18], and their potential for ESCSS detection is rarely investigated.

The widespread use of high-throughput techniques in ESCC allowed us to view the landscape of genomic features of this disease. DNA methylation as one type of epigenetic modification has received the most attention. Recent findings suggested that aberrant DNA methylation can be used as a signal for the early detection of ESCC. For example, hypermethylated *CDKN2A, CDKN2B*, and *TFF1* were found in the early stages of ESCC [25]. In a study combining 850k and 450k methylation data in ESCC, DNA methylation was more frequent and robust in tumor tissues than normal mucosa, with 1/4 of the hypermethylated genes (165 genes) being observed in early ESCC (stage I-II) [26]. Investigations based on whole-genome bisulfite sequencing (WGBS) data revealed that about 2% (∼36,000) of CpG sites were in hypermethylated status, including inactivated negative regulators of the Wnt pathway due to aberrant methylation [27]. These pioneering studies provide us a treasure trove to identify more effective biomarkers for the early detection of ESCC.

Although many studies have revealed aberrant DNA methylation in ESCC, most have focused on only a tiny fraction of genome. The widely used 450k methylation microarray data cover approximately 480,000 CpG sites [28], representing the local genomic methylation status. Therefore, it is necessary to explore the methylation features from a global perspective. In this study, we investigated the methylation patterns of ESCC in genome-wide using WGBS data and elaborated the possible biological functions of these aberrantly methylated genes. Finally, the ability to detect ESCC and nine other cancer types was shown by integrating multiple datasets and screening several methylation genes as potential markers.

## Methods

### Data preparation and preprocessing

GSE149608 [27] and GSE52826 [29] datasets were retrieved from the GEO database (https://www.ncbi.nlm.nih.gov/geo/) as both consisted of tumor and paired normal samples. The two datasets were generated by whole-genome bisulfite sequencing (WGBS) and Illumina HumanMethylation450k, covering over 18 million and 480,000 CpG sites. The methylation value of each CpG site was represented by the percentage of methylated reads in total reads that covered this site for WGBS data and by the signal intensity value of β (range: 0-1) for 450k microarray data. TCGAbiolinks tools [30] was used to download the level 3 methylation data, the corresponding clinical features of ESCC, and paired normal samples from The Cancer Genome Atlas (TCGA) Program (https://portal.gdc.cancer.gov/). CpG sites were removed during the data preprocessing if the methylation values were 0 in all samples of WGBS dataset or the β values were missing in more than 90% of the samples in 450k dataset. Then, KNN algorithm was used to fill the missing values.

The preprocessed datasets were shown in **Table 1**, containing 29 normal samples and 109 tumor samples. The clinical information of GSE149608 was collected from GEO database simultaneously but was not consistent with the reference paper. Here, we determined their exact clinical characteristics (**Supplementary table 1**) according to the information provided by the reference, which included two early-stage patients (patient3 and patient10), five intermediate-stage patients (patient2, patient5-8), and two late-stage patients (patient1 and patient4) stages. All cases in GSE52826 dataset were early-stage ESCC (**Supplementary table 2**).

**Table 1.** The three eligible datasets used in this study.

| Dataset | Normal (n) | Tumor (n) | Platform |
| --- | --- | --- | --- |
| GSE149608 | 9 | 9 | WGBS |
| GSE52826 | 4 | 4 | Illumina HumanMethylation450k |
| TCGA ESCC | 16 | 96 | Illumina HumanMethylation450k |

### Human tissue and blood samples

Twenty formalin-fixed paraffin-embedded esophageal squamous carcinoma and adjacent normal sample were collected from the department of pathology of the First Affiliated Hospital of Zhengzhou University in the study. Meanwhile, twenty healthy blood samples were obtained from the blood transfusion department, where the residual blood samples were collected from healthy individuals after blood donation. The healthy blood samples were included because they represented the methylation status of candidate targets on healthy individuals and will also facilitate investigating their potential role as blood diagnostic markers. Twenty plasma samples were also collected from ESCC patients, but only 13 samples obtained adequate cell-free DNA amounts. Clinical features of the samples are displayed in **Supplementary table 3**. Referring to the NCCN guidelines, in this study, we defined Tis (high-grade dysplasia), T1 (T1a and T1b), and some T2 (without lymph node metastasis and distal metastasis) as early-stage ESCC. All individual identifiers have been removed. The Ethics Committee of the First Affiliated Hospital of Zhengzhou University approved this study (approval number 2020-KY-0152).

### Identification of differentially methylated genes

Since the vast majority of CpG sites in the whole genome were covered in WGBS data represented the comprehensive epigenetic information, the GSE149608 dataset was selected to identify differentially methylated CpG sites (DMC) between tumor and paired normal samples using paired student t-test, with P-value <0.05 and fold change ≥2 as the significance threshold. We defined hypermethylated DMGs in normal and tumor samples as NC-DMCs and ESCC-DMCs respectively. The methylation status of adjacent CpG sites are usually highly coupled with each other, and they tend to be located in a small genomic region [31], therefore we further identified the differentially methylated regions (DMR) based on DMCs using a modified sliding window approach described in our previous study [32]. Briefly, to facilitate developing methylation specific PCR (MSP) assay, the maximum length of DMRs was set to 100 bp, and each DMR contained at least three DMCs, and the distance between two adjacent DMCs was less than 50bp. NC-DMRs and ESCC-DMRs were then identified separately, and the genes covered by NC- or ESCC-DMRs were defined as differentially methylated genes (NC-DMG or ESCC-DMG), except for those overlapped with both NC- and ESCC-DMRs. Since ESCC-DMRs are more eligible for MSP, only ESCC-DMGs were selected for subsequent analysis.

### Function enrichment analysis

Gene ontology (GO) and KEGG pathway enrichment analysis was performed using the ‘clusterProfiler’ R package for NC-DMGs and ESCC-DMGs. All human genes were used as the background with q-value < 0.05 as the significance threshold. Gene family enrichment analysis was implemented by integrating gene family terms provided by HGNC (HUGO Gene Nomenclature Committee) [33] and the chi-square test. Briefly, the human gene family data were downloaded from HGNC (www.genenames.org), which records the human genes and their corresponding families. For each family, we counted the number of DMGs belonging to this family. The overrepresented gene families were assessed with all human genes and DMGs as background. The chi-square test *P-value*< 0.05 was selected as the significance threshold to determine enriched families for NC-DMGs and ESCC-DMGs.

### DNA extraction and BS-treatment

DNA of tissue and blood samples were extracted using paraffin-embedded tissue DNA Rapid Extraction Kit (TIANGEN^®^, Beijing) and TIANamp Genomic DNA Kit (TIANGEN^®^, Beijing), respectively. BS-treatment and purification for the extracted DNA were subsequently carried out using the Nucleic Acid Purification Kit (Ammunition^®^, Wuhan). The basic principle of BS-treatment is that unmethylated cytosines in denatured DNA can be converted to uracil by bisulfite ions, while methylated cytosines are remained. The methylation status then can be determined by methylation-specific PCR.

### Methylation-specific PCR and Sanger sequencing

Methylation-specific PCR experiments and Sanger sequencing were performed to determine the methylation status of candidate targets. Two genes, *HOXD1* and *GSX1*, were selected for MSP because they showed good abilities to discriminate cancer samples from normal samples. Besides, high densities of differentially methylated CpG sites were found within the two genes, which allowed us to design appropriate MSP primers. The designed primers for *HOXD1* and *GSX1* were listed in **Table 2**. The amplified regions covered DMR4, DMR5, and DMR6 of *HOXD1*, and DMR16 and DMR17 of *GSX1* (**Supplementary table 4**). The fully unmethylated and methylated DNA fragments for the two targets were synthesized as negative and positive controls. The MSP amplification system was 20ul, including 7ul of dd-water, 10ul of 2 ×T5 Fast qPCR Mix (SYBRGreenI), 0.5ul of forward and reverse primers (10 uM), and 2ul of BS-converted DNA. PCR reaction was pre-denaturation at 95°C for 1 min, denaturation at 95°C for 10 s, annealing and extension at 60°C for 45s. The melt curve reaction was 95°C ∼ 15s, 60°C ∼ 1 min, and 95°C ∼ 15s. Quantitative real-time PCR was performed on a 7500 device (Thermo Fisher, USA). The MSP products were then used for Sanger sequencing. After BS-treatment, the methylated CpG site would remain C, and the un-methylated C would be T. Therefore, the methylation status of target CpG sites can be determined according to the result of Sanger sequencing.

**Table 2.**
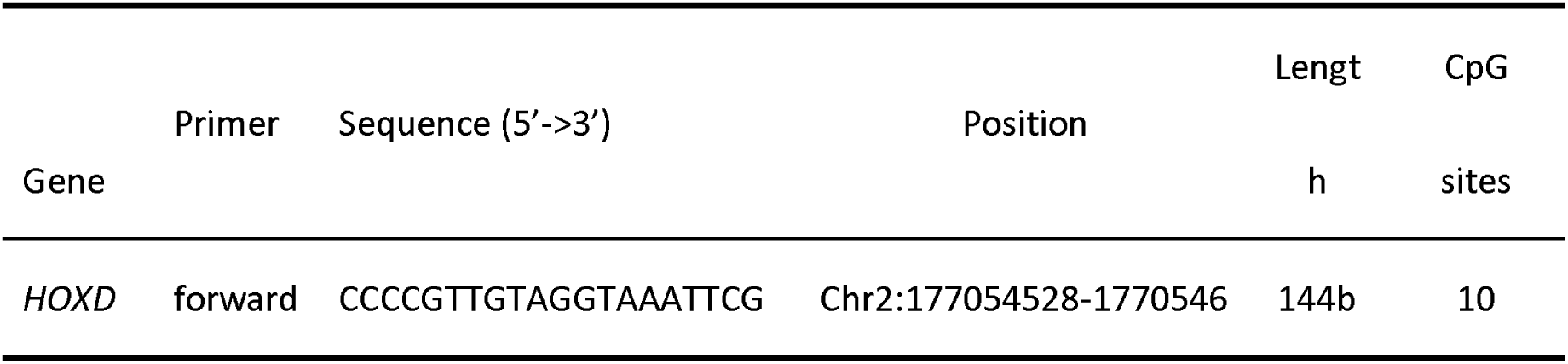
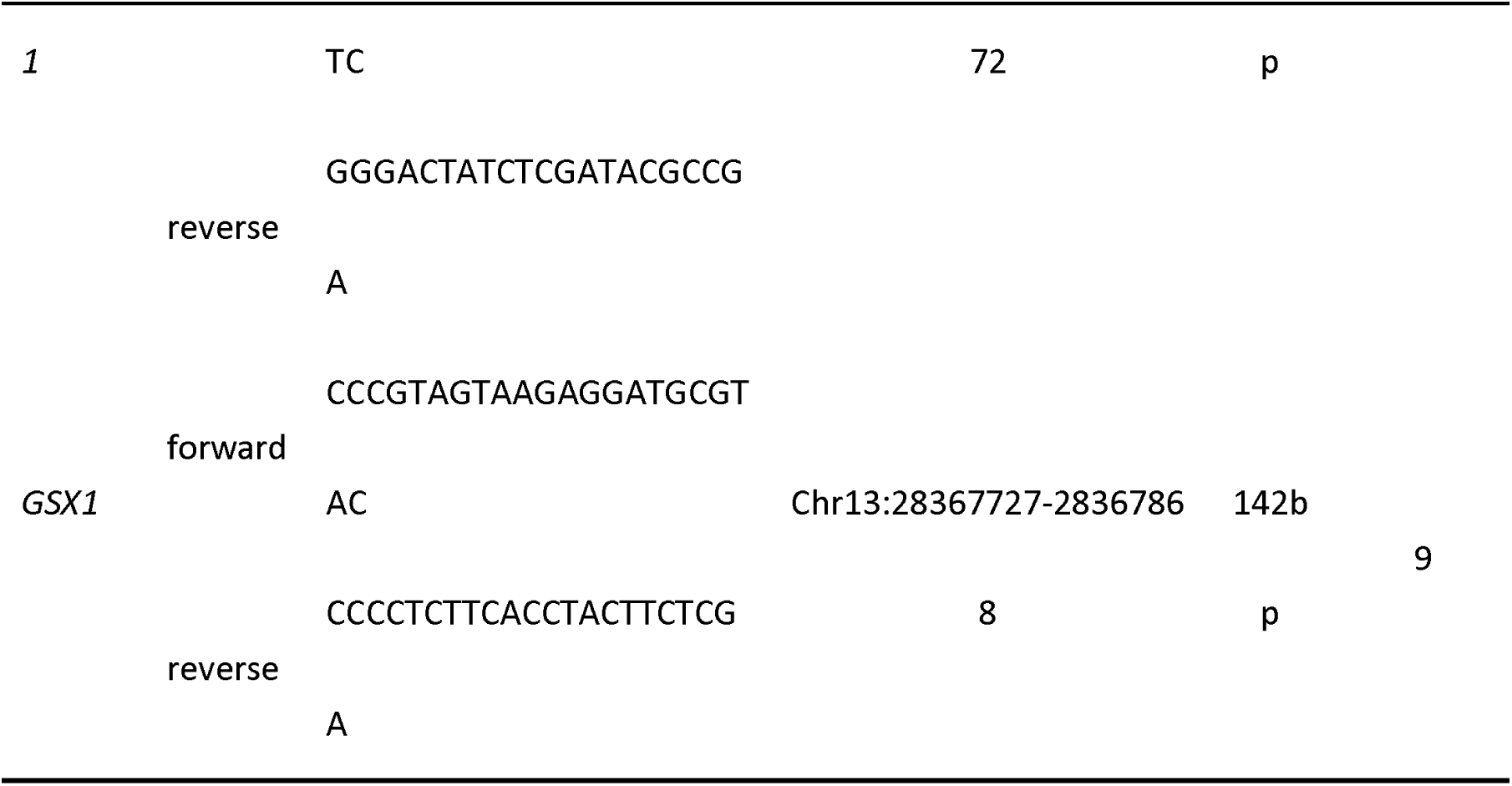
Primers used for methylation-specific PCR.

### ROC analysis

For the candidate target genes, we view the sample types (normal or tumor) as the response variable of their methylation values to develop a classifier for ESCC using logistic regression. The logistic probability, herein defined as risk scores for samples, was then estimated. Typically, samples would be recognized as positive if the risk scores were > 0.5 and vice versa as negative. This study used ROC curve to determine the appropriate threshold and AUC value to assess the classifier performance. The optimal threshold was locked when Youden index reached the maximum. Samples were subsequently divided into a positive group if the scores were higher than this threshold or a negative group if the scores were less than this threshold. All samples were classified into four categories, true positive (TP), true negative (TN), false positive (FP), and false negative (FN) according to their sample types and predicted types. The sensitivity and specificity were calculated using the following formulas:

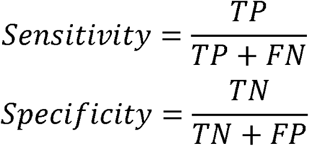

### Statistical analysis

Data preprocessing, statistical analysis, and other analysis in this study were implemented in R software (v3.6.1). For continuous variables, paired student t-test was used for comparisons of paired normal and tumor samples, and the wilcoxon rank-sum test for unpaired datasets. The Kruskal-Wallis test was performed for comparisons of multiple groups. For category variables, chi-square test was used to estimate the difference between groups. Logistic regression was implemented using the R package ‘glmnet’ with the parameter ‘family=binomial’. ROC analysis was conducted using the package ‘pROC’ [34] with default parameters.

## Results

### Landscape of the aberrantly methylated CpGs between normal and ESCC

The flowchart of this study was presented in **Figure 1A**. The following analyses were performed to survey the landscape of ESCC methylome. Firstly, differentially methylated CpG sites between normal and tumor samples were identified using WGBS data. Due to the highly coordinated CpG sites being often tightly coupled with each other, we further identified differentially methylated regions (DMR) by a modified sliding window method. Differentially methylated genes (DMG) were then identified based on the genomic coordinates of DMRs and genes (see the method). Here we defined DMR or DMG as NC-DMR and NC-DMG if their methylation levels were higher on normal controls than tumor samples and vice versa as ESCC-DMR and ESCC-DMG. GO and KEGG pathway enrichment analyses were conducted to explore the potential functions of these DMGs. We also performed gene family analysis to identify significantly enriched gene families. The HOXL subclass homeobox family was selected to validate on two independent datasets and by Sanger sequencing in our custom samples. We further investigated the potential utility of the family genes as markers for cancer detection.

**Figure 1.**
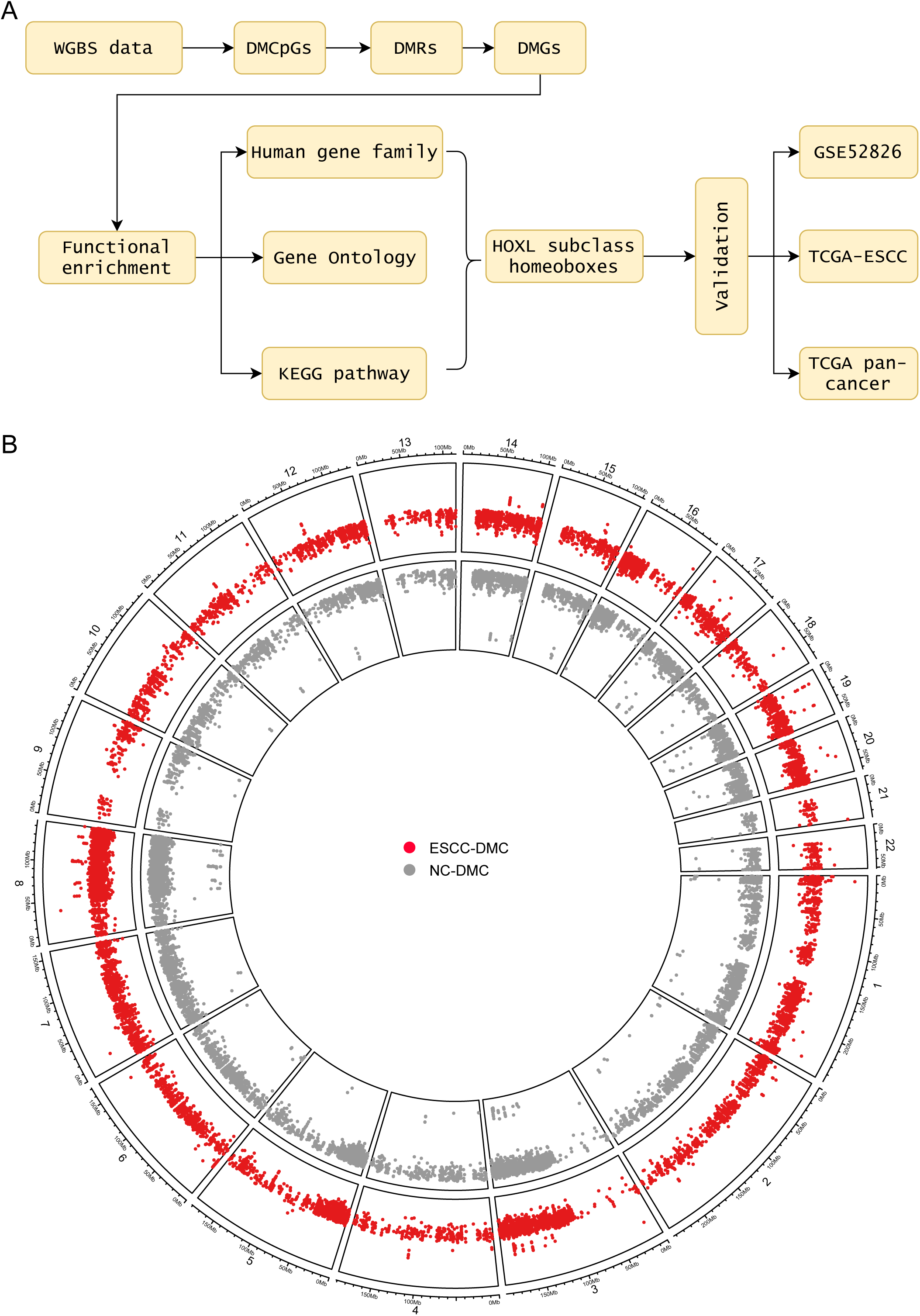
Methylation profiling of ESCC and paired normal samples. **A:** An overview of this study. TCGA pan-cancer included the 9 most common cancer types: esophageal carcinoma (ESCA), esophageal squamous cell carcinoma (ESCC), stomach adenocarcinoma (STAD), colorectal cancer (CRC), liver hepatocellular carcinoma (LIHC), pancreatic adenocarcinoma (PAAD), breast invasive carcinoma (BRCA), lung adenocarcinoma (LUAD), lung squamous cell carcinoma (LUSC). **B:** The identified DMCs across the whole genome. The red and gray points represented the ESCC-DMCs and NC-DMCs respectively. Chromosome X and Y were excluded in **Figure 1B**. WGBS: whole-genome bisulfite sequencing, DMC: differentially methylated CpG, DMR: differentially methylated region, DMG: differentially methylated gene.

We identified 438558 DMCs, including 361341 NC-DMCs and 69278 ESCC-DMCs (**Figure 1B, supplementary table 5**). Chromosome 8 has the most NC-DMCs, while the most ESCC-DMCs were in chromosomes 1 and 2 (**Supplementary figure 1A**). The distance between ESCC-DMCs was smaller than that of NC-DMCs (**Supplementary figure 1B**). Further investigations indicated that the methylation levels between adjacent DMCs were strongly correlated, and the correlation coefficients of adjacent ESCC-DMCs were much higher than NC-DMCs (0.73 vs. 0.70, *P* < 0.05, **Supplementary figure 1C**).

### Identification of DMGs

We obtained 6422 NC-DMRs and 9040 ESCC-DMRs based on the DMCs. Similarly, most NC-DMRs were found in chromosome 8, while the majority of ESCC-DMRs were in chromosome 1 and 2 (**Supplementary figure 2A**). A smaller distance was also observed for adjacent ESCC-DMRs than that of NC-DMRs (**Supplementary figure 2B**). The average DMC count of ESCC-DMRs was higher than that of NC-DMRs (**Figure 2A**), and ESCC-DMRs tended to be located in inner-genic regions, whereas NC-DMRs more often located in intergenic regions (**Figure 2B**). According to the genomic coordinates of DMRs and genes, we identified 733 NC-DMGs and 906 ESCC-DMGs, of which 71 genes were shared by both (**Supplementary table 6**). Both NC- and ESCC-DMGs were mostly distributed in chromosome 19, which was different from DMCs and DMRs. The methylation values were able to separate ESCC-DMGs and NC-DMGs clearly, and ESCC-DMGs showed more concentrated than NC-DMGs (**Figure 2C**, 1st-3rd quantile: [29.32-43.64] vs. [13.86-34.14]). Functional enrichment analysis revealed that neuroactive ligand-receptor interaction was the most significantly enriched pathway for NC-DMGs (**Figure 2D**). For ESCC-DMGs, calcium signaling pathway was the most significantly enriched pathway (**Figure 2E**). The term of CD molecules was the major enriched gene family for NC-DMGs, whereas NKL subclass (**Figure 2F, Supplementary table 7**), HOXL subclass, and Zinc fingers C2H2-type were the 3 top-ranked enriched families for ESCC-DMGs (**Figure 2G, Supplementary table 8**).

**Figure 2.**
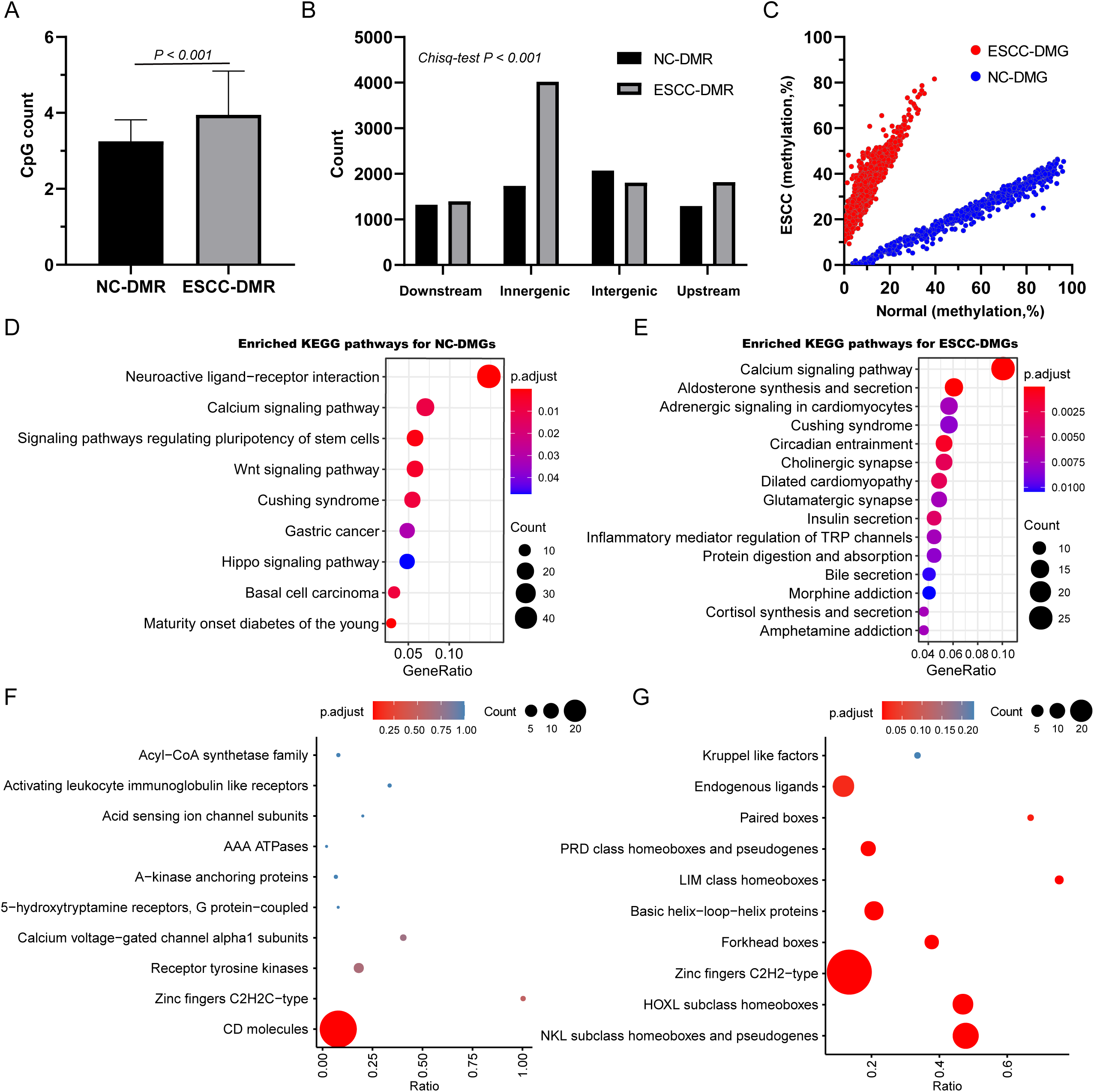
Functional enrichment analysis of differentially methylated genes. **A:** The number of DMCs in ESCC-DMRs and NC-DMRs. **B:** The genome distribution of ESCC-DMRs and NC-DMRs. **C:** The methylation levels of DMGs in normal and ESCC samples. **D&E:** The enriched KEGG pathways of NC-DMGs **(D)** and ESCC-DMGs **(E). F&G:** The top 10 enriched gene families of NC-DMGs **(F)** and ESCC-DMGs **(D)**. The point size in **Figure 2D-F** represented the mapped gene numbers, and color indicated the enriched p value.

### The aberrant methylation of HOXL subclass homeoboxes

We selected the second-ranked HOXL subclass homeobox family for further analysis, as more than half (n=24, 66.67%) of the genes were identified as ESCC-DMGs. K-means clustering showed that the 36 genes could be clustered into 3 groups (**Figure 3A**). Group 1 consisted of 11 genes that were hypermethylated on both normal and ESCC samples. Twenty-four genes, all of which were ESCC-DMGs, were in group 2. Furthermore, group 2 genes were divided into three sub-groups, subgroup 1 (n=10), subgroup 2 (n=12), and subgroup 3 (n=2). Only one gene, *HOXA7*, was in group 3 and showed low methylation levels on normal and ESCC samples. Overall, genes of subgroup 2 showed the lowest methylation levels on normal samples than sub group 1 and sub group 3 (**Figure 3B**). Correlation analysis revealed significant positive correlations between group 2 genes, except for *MNX1* and *GBX1* (**Figure 3C**).

**Figure 3.**
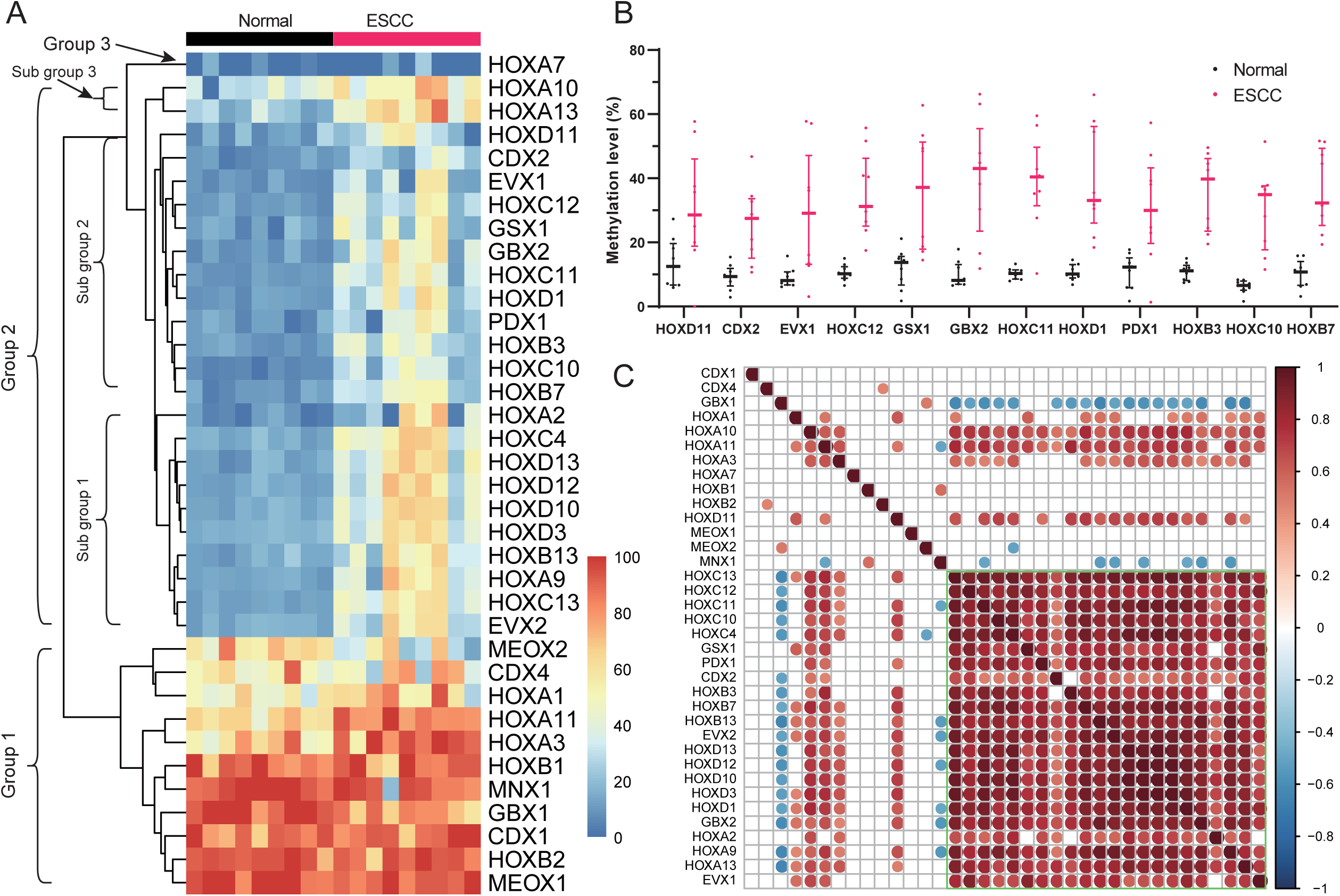
Methylation patterns of HOXL subclass homeoboxes. **A:** The methylation levels of HOXL subclass homeoboxes genes in normal and ESCC samples. **B:** The methylation levels of subgroup 2 genes between normal and ESCC samples. **C:** The methylation correlation of HOXL subclass homeobox genes. The correlation coefficients were estimated using Pearson’s method. The β values of each gene in **Figure 3A-C** were calculated using TCGA ESCC dataset.

### Validation of sub-group 2 genes of HOXL subclass homeobox

Using GSE52826 dataset, we verified the methylation characteristics of subgroup 2 genes (n=10) in ESCC and paired normal samples (**Figure 4A**). After the exclusion of outlier samples (GSM1276746 and matched GSM1276750) (**Supplementary figure 3**), six genes were validated hypermethylated in tumor samples (**Figure 4B**). Since *HOXC12* showed high methylation levels in normal samples, we further investigated methylation patterns of the rest five genes on TCGA ESCC dataset (**Figure 4C**). The methylation of *HOXC11, HOXC10*, and *HOXD1* in normal samples were lower than *GSX1* and *CDX2* (**Figure 4D**), suggesting their low methylation background. ROC analysis indicated that methylation values of *HOXC10* and *HOXD1* showed the best performance in discriminating ESCC from normal samples, with both AUC reached 0.85 (**Figure 4E**). Further investigations revealed that four DMRs located in *HOXC10*, and HOXC10-R3 presented the largest delta methylation value (**Figure 4F**). *HOXD1* contained seven DMRs, with HOXD1-R4 showed the largest delta methylation value (**Figure 4G**). When stratified TCGA ESCCs by different pathological stages, both genes showed hypermethylated across all stages, including stages IA and IB (**supplementary figure 4**).

**Figure 4.**
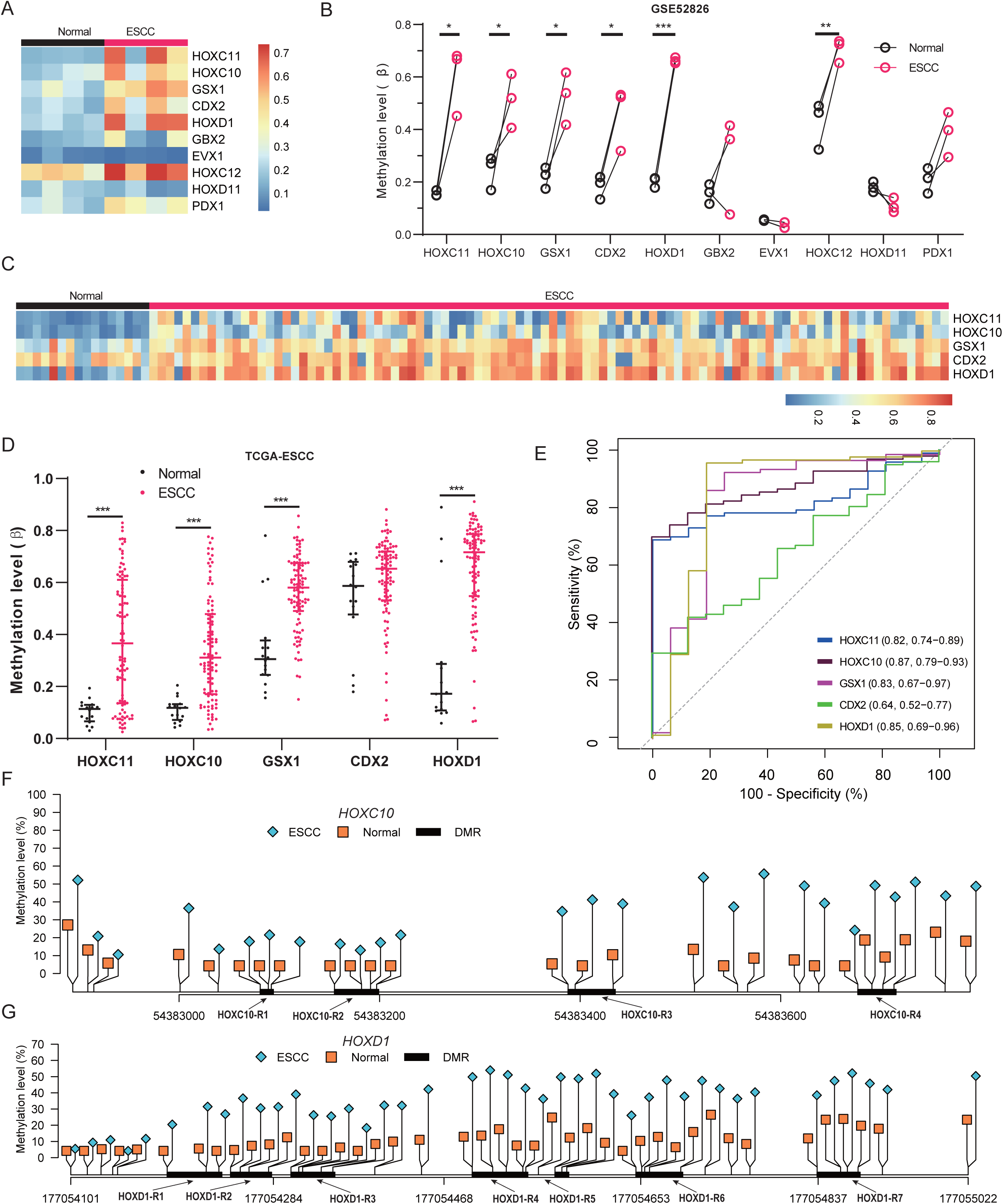
Methylation patterns of sub-group 2 genes in validation sets. A: The methylation levels of sub-group 2 genes in normal and ESCC samples in GSE52826 dataset. **B:** The differentially methylated six genes between ESCC and paired normal samples. Paired student t-test was used to estimate significant p values. **C:** The methylation levels of five genes in normal and ESCC samples in TCGA-ESCC. **D:** The differentially methylated four genes between ESCC and normal samples. The Wilcoxon rank sum test was used to estimate significant p values. **E:** ROC curves of the four genes in classifying TCGA-ESCC and normal samples. **F-G:** The identified ESCC-DMRs of *HOXC10* and *HOXD1* represented by R1-R4 and R1-R7 respectively.

### The performance of HOX-score for ESCC classification

We used logistic regression to develop an ESCC classification model based on HOXC10/HOXD1 methylation using the TCGA ESCC cohort. The risk score, defined as HOXL-score, was then estimated for each sample. The normal samples showed the lowest risk scores than ESCC and ESCA (**Figure 5A, median: 0.28, 0.88, and 0.99**). According to the mechanism of logistic regression, the HOXL score represented the probability that a sample was classified as cancer. Therefore, we attempted to classify ESCC and normal samples using HOXL scores. ROC curve analysis indicated that the AUC reached 0.96 (95% CI: 0.91-0.99) for ESCC (**Figure 5B**). The optimal threshold determined by the Youden index was 0.72, with a sensitivity of 94.8% and specificity of 87.5%. Besides, for ESCA, the AUC was 0.83 (95% CI: 0.72-0.93), with sensitivity and specificity of 83.1% and 87.5% at the optimal threshold (**Figure 5B**). No significant variations were observed for HOXL-score in detecting both ESCC and ESCA stratified by gender, age, and stage at the threshold of 0.72 (**Table 3**).

**Figure 5.**
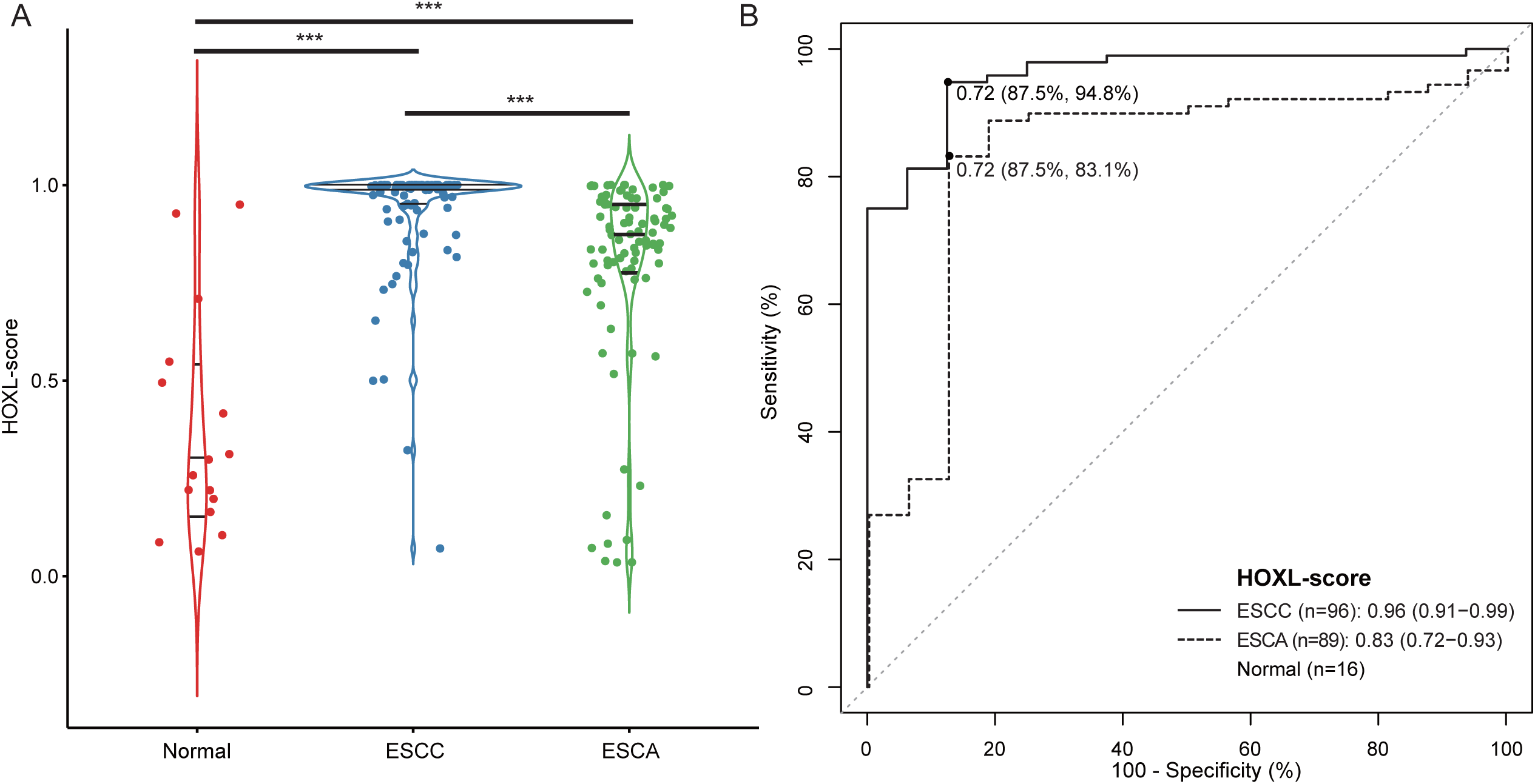
The performance of HOXL score for ESCC classification. **A:** The HOXL scores of normal, ESCC and ESCA samples. For two group comparisons, the Wilcoxon rank sum test was used. For three group comparison, the Kruskal test was used. The three lines from top to bottom in each category indicated the median and 90th percentile of scores. **B:** ROC curves of HOXL-score for ESCC and ESCA classification. The points indicated the best cutoff of HOXL-scores, and the percentages were the best specificity and sensitivity estimated by Youden’s index.

**Table 3.**
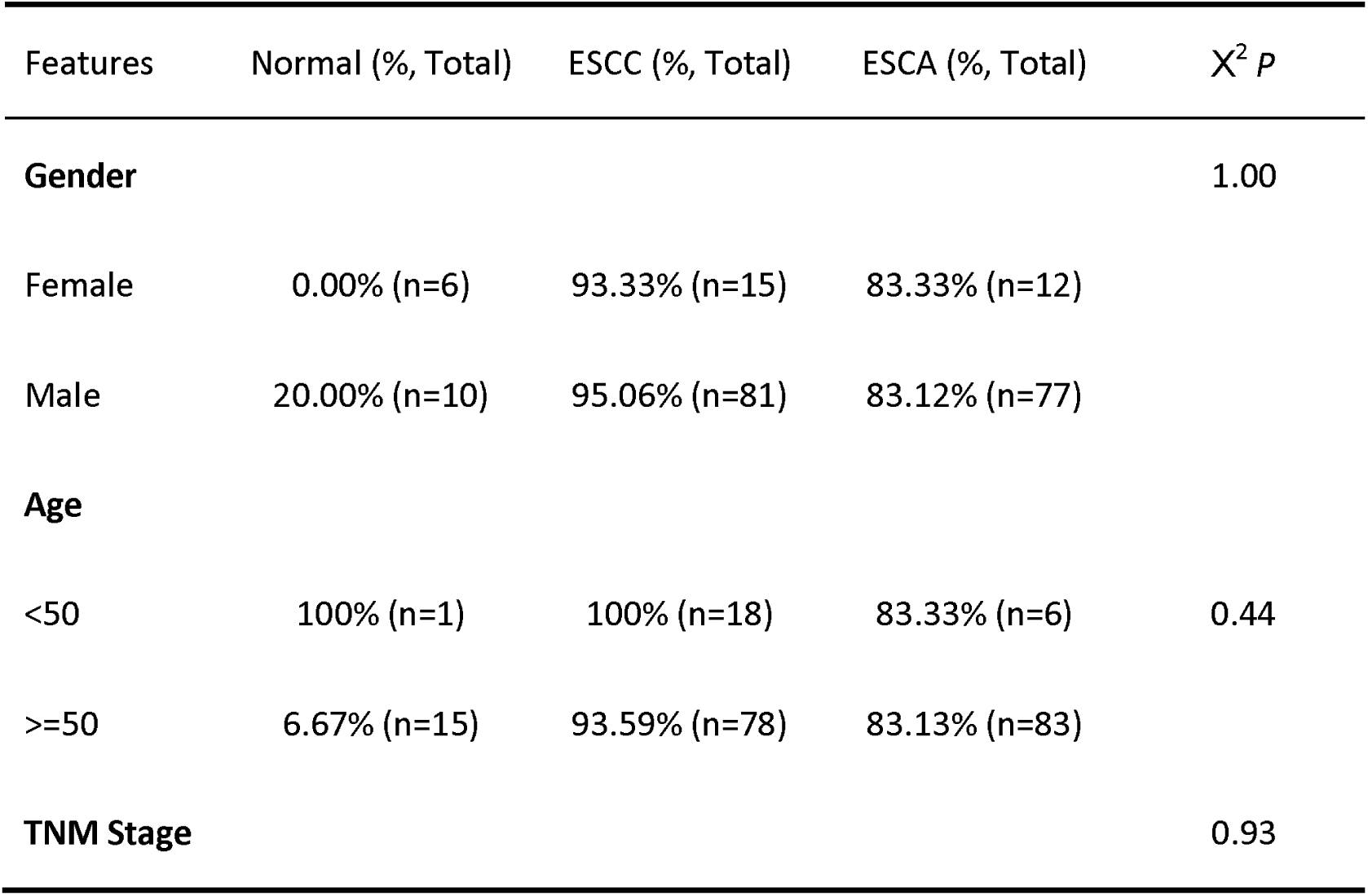
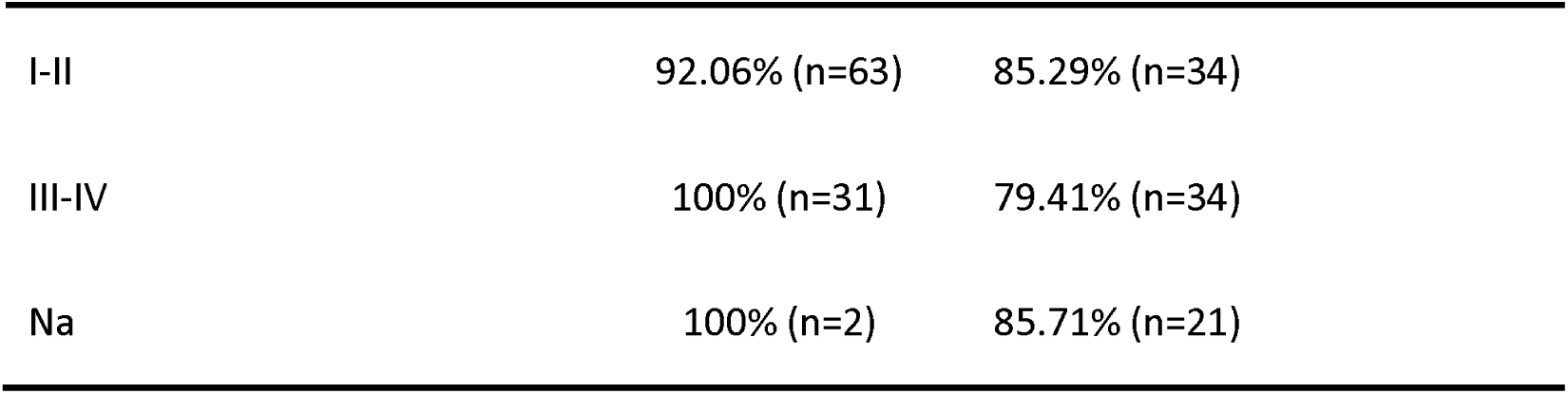
The positive detection rates of HOXL-score for ESCC and ESCA with different clinical features.

The diagnostic performance of 13 potential methylation markers, *CDKN2A, CDKN2B, TFF1, MGMT, MLH1, DAPK1, SCGB3A1, TFPI2, DACH1, SOX17, CHFR, CDH1, APC* that have been reported in ESCC were also evaluated using the same approach in TCGA ESCC dataset (**supplementary figure 5**). We observed the highest AUC value (0.96 [95%CI: 0.91-0.98]) for *DAPK1* methylation, which is comparable to the HOXL-score. The AUC values of *CDKN2A* and *TFF1* were 0.85, which equaled *HOXD1* but lower than *HOXC10*. These results suggested that the two genes and their combination showed great potential for ESCC detection.

### The methylation patterns of HOXL subclass homeoboxes in pan-cancers

We observed extensive hypermethylation of HOXL subclass homeobox genes on ESCC and then investigated their methylation patterns on nine other cancer types, including STAD (n=393), COAD/READ (n=379), LIHC (n=374), PAAD (n=183), PRAD (n=495), BRCA (n=778), LUAD (n=456) and LUSC (n=364). Forty (83.33%) and eight genes (16.67%) were found hypermethylated on cancer and normal samples, respectively. *HOXD10* showed the highest delta β between normal and tumor samples, with widespread hypermethylated on most cancer types, except for LIHC (**Figure 6A**). The similarity was observed for *HOXD12, HOXD1*, and *HOXC5* (**Supplementary table 9**). *GSX1* had the smallest standard error (sd) of β values on both cancer and normal samples, indicating its good uniformity across the ten cancer types (**Figure 6B**). Therefore, we further evaluated the potential utility of *GSX1* methylation in detecting pan-cancers. Pan-cancer samples were randomly divided into training and testing sets by 1:1. AUC was 0.89 (95% CI: 0.87-0.91) for training set with a sensitivity and specificity of 84.1% and 85.4% at the optimal threshold of 0.88. For the testing set, the AUC was 0.88 (95% CI: 0.86-0.90), and the sensitivity and specificity were 84.2% and 82.7%, respectively (**Figure 6C**). In addition, *HOXC10* showed the highest methylation levels on ESCC than other cancer types and normal samples (**Figure 6D**), suggesting that it could be used as a marker specifical for ESCC.

**Figure 6.**
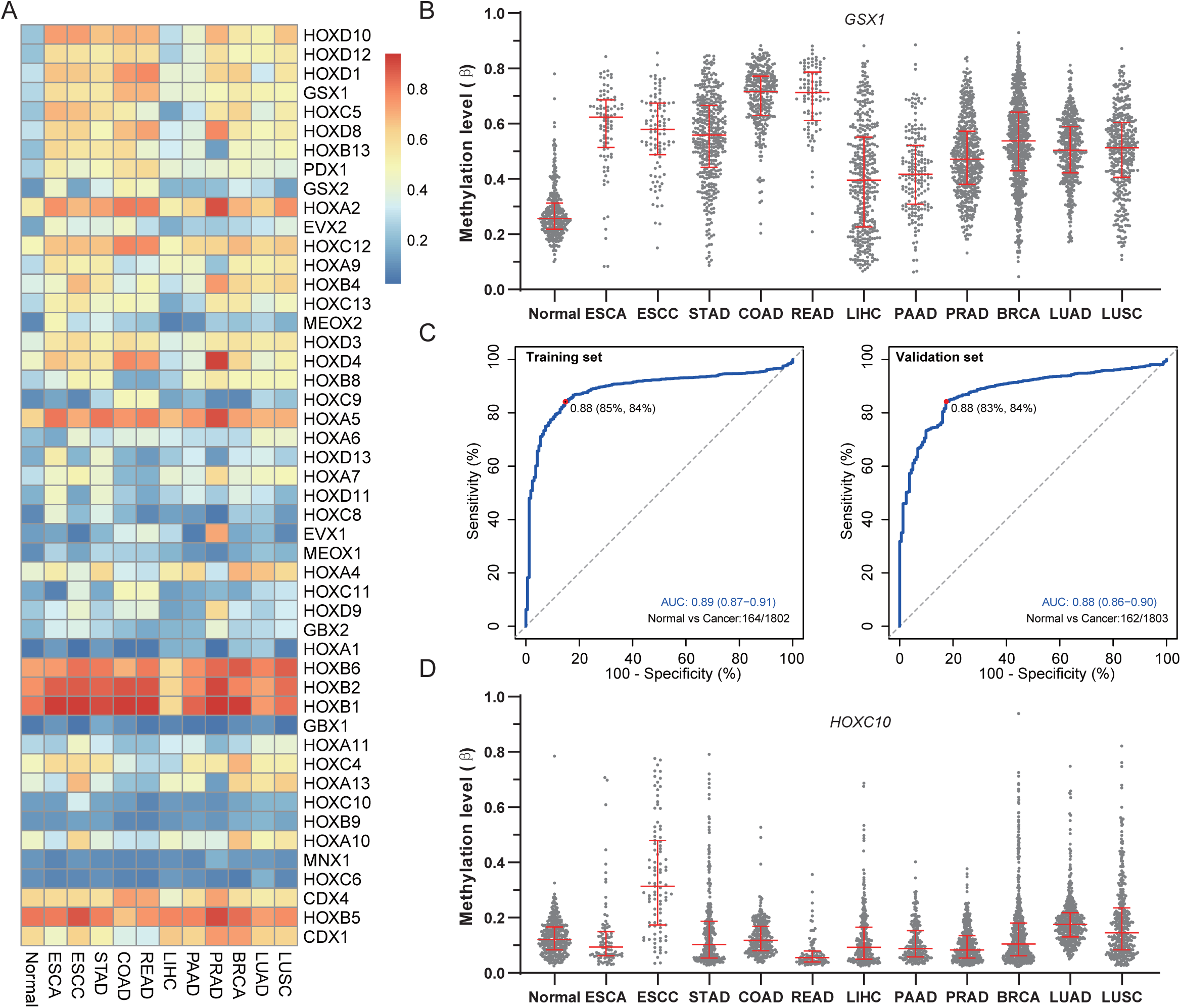
The potential of HOXL subclass homeoboxes for pan-cancers detection. **A:** The methylation levels of HOXL subclass homeoboxes genes in normal samples and 9 cancer types. The gene methylation level of each category was represented by the average β values of all samples in each cancer. **B:** The β values of *GSX1* in normal samples and multiple cancer types. **C:** ROC curves showed the classification performance of *GSX1* methylation in training and test sets. The red points indicated the optimal cutoff of probabilities estimated by logistic regression. **D:** The β values of *HOXC10* in normal samples and multiple cancer types. For panel **B** and **D**, the red lines represented the median and 90th percentile β values.

### Validation of the methylation status of *HOXD1* and *GSX1* by sequencing

Among all collected samples, amplification products of *HOXD1* were obtained from 4 normal samples, 19 ESCC tissues and 19 blood samples. For *GSX1*, products were obtained from 14 normal samples, 19 cancerous tissues and 19 blood samples. The results of Sanger sequencing indicated that methylation events of the target regions on both genes occurred more frequently on ESCC samples (**Table 4**). For all CpG sites, the methylation frequency on ESCC samples was significantly higher than that on normal samples (**Supplementary table 10**). Sanger sequencing was also conducted for 13 cfDNA samples collected from ESCC patients’ plasma. In the 13 cfDNA samples, we observed that 90.77% (118/130) of *HOXD1* CpG sites were methylated, and 92.31% (108/117) of *GSX1* CpG sites were methylated (**Figure 7A&B, Table 4**). The methylation ratios of cfDNA samples were a little lower than tissue samples (99.47% for *HOXD1* and 99.42% for *GSX1*), which might be attributed to the low cfDNA amount in plasma resulting in failed detection. Besides, we observed lower methylation levels of *HOXD1* than *GSX1* on normal samples in TCGA dataset, which was also revealed by the sequencing results that fewer methylation events occurred on normal samples for *HOXD1* than *GSX1* (**Figure 7A&B**).

**Table 4.**
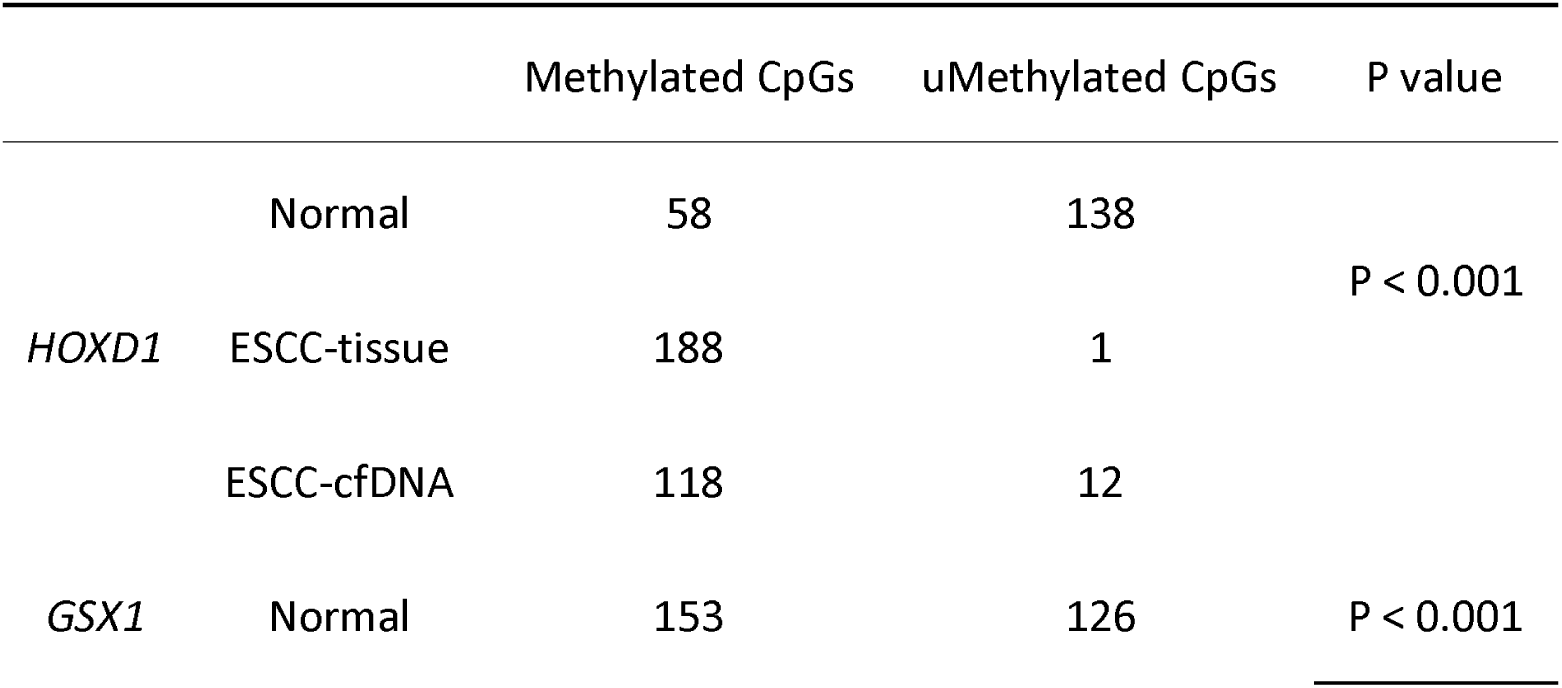
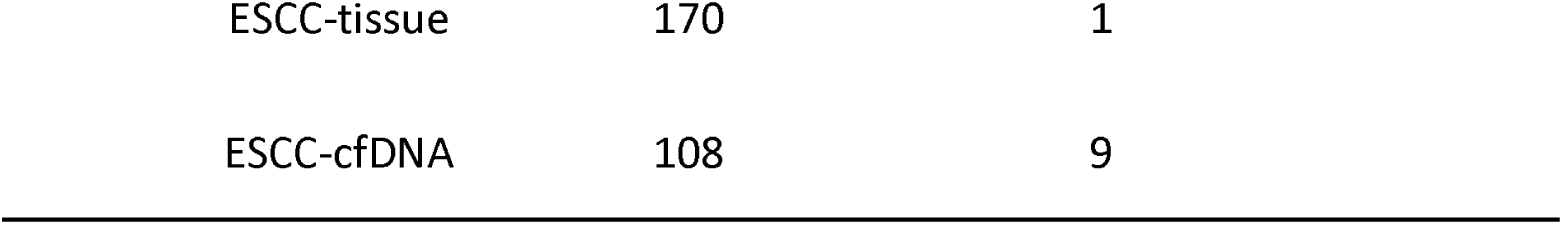
The methylation frequency of the CpG sites in target regions.

**Figure 7.**
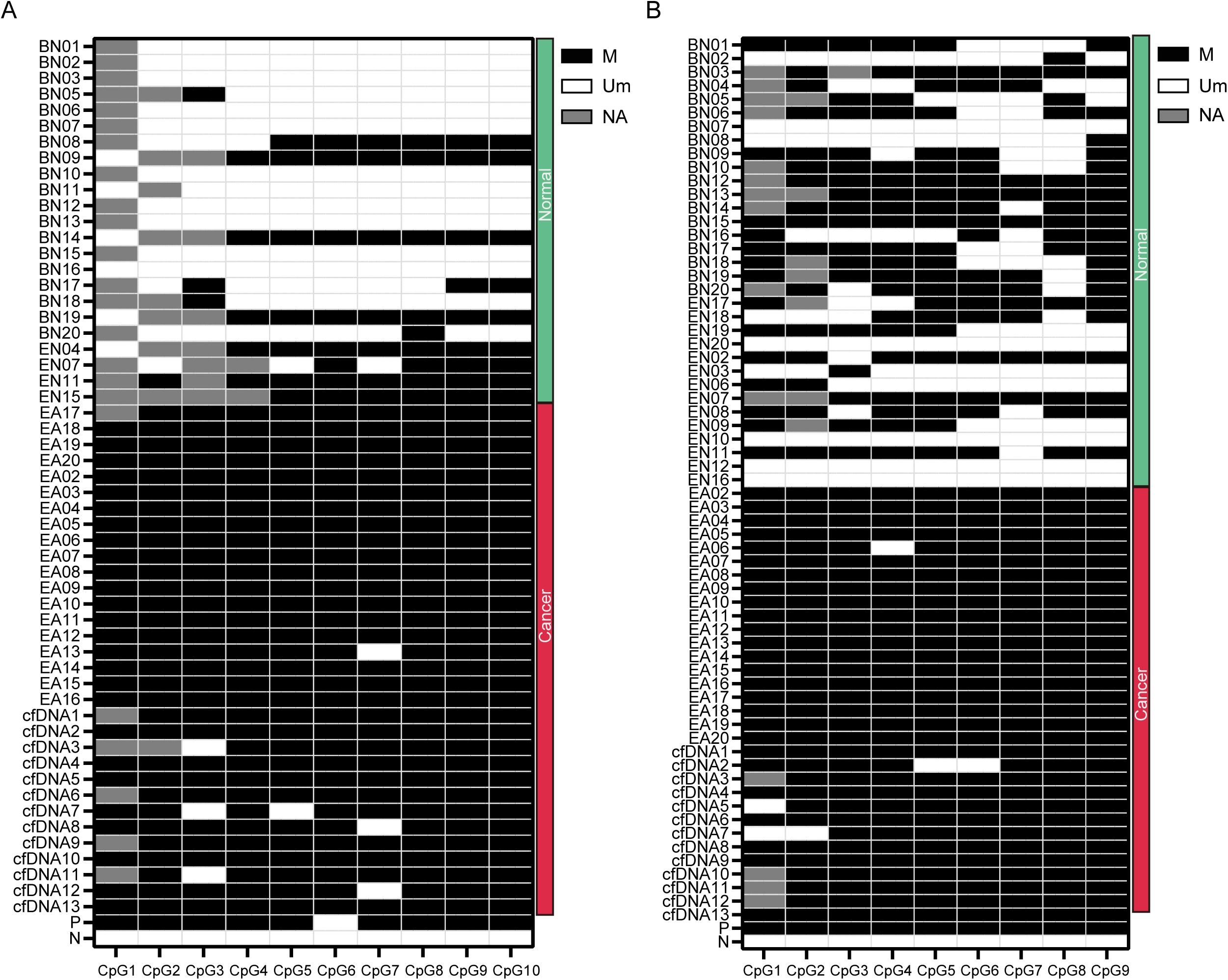
The methylation status of *HOXD1* (A) and *GSX1* (B) between normal, ESCC-tissue and -plasma samples. Normal and ESCC samples were indicated by green and red side bars. BN, EN, EA, and cfDNA represented the blood samples, adjacent normal samples, ESCC-tissue and plasma samples, respectively. The methylated and unmethylated status of each CpG site were presented by black and white color in the heatmap. P and N indicated the positive and negative controls.

## Discussions

Abnormal DNA methylation is the most common epigenetic variation in human diseases and has been observed in various cancer types, including esophageal cancer. Therefore, further investigations of DNA methylation will facilitate understanding its role in tumor formation and developing more effective methylation-based biomarkers for cancer early detection. This study identified more than 40,000 differentially methylated CpG sites between ESCC and paired normal samples using WGBS data. A sliding window method was adopted to determine the differentially methylated regions and differentially methylated genes involved in various biological processes. We further observed the frequent hypermethylation of HOXL subclass homeoboxes genes on ESCC, and the subsequent studies suggested their potential utility in discriminating ESCC and nine other cancer types from normal samples.

We identified 369,119 NC-DMCs and 69,439 ESCC-DMCs from the WGBS data covering 18 million CpGs, accounting for 2.44% of the total CpG sites, close to the previously reported 2.7% of methylated CpG sites [27]. Previous studies based on LINE-1, an alternative indicator of the whole genome, consistently observed genome-wide hypomethylation in ESCC [35]. Similar results were obtained in this study, with more NC-DMCs identified than ESCC-DMCs (approximately five times), suggesting the prevalent hypomethylation events in ESCC. The distance between two adjacent ESCC-DMCs was smaller than that of two adjacent NC-DMCs, indicating that the ESCC-DMCs preferred to be more concentrated in one region. This feature was also evidenced by the fact that more ESCC-DMRs were identified (NC-DMRs/ESCC-DMRs: 6422/9040). The subsequently identified 733 NC-DMGs and 906 ESCC-DMGs included *PAX1* [36] and *STK3* [37], of which both have been attempted as diagnostic markers for ESCC. Functional enrichment results revealed that ESCC-DMGs enriched in multiple biological processes, such as the reported cell cycle regulation and Wnt signaling pathways [35]. Notably, the top 3 enriched families, NKL subclass homeoboxes and pseudogenes, HOXL subclass homeoboxes and Zinc fingers, were frequently observed hypermethylated in ESCC. A couple of studies have revealed that the Zinc fingers family genes, such as *ZNF382, ZNF582, ZNF418*, and *ZNF542*, are methylated on ESCC [36-38]. However, the NKL and HOXL subclass, both of which belong to the homeoboxes superfamily, were not well reported. Our findings demonstrated the hypermethylation events of homeoboxes superfamily genes on ESCC, suggesting their potential roles in esophageal tumorigenesis.

This study revealed the widespread hypermethylation of HOXL subclass homeobox genes on ESCC in multiple independent datasets. Methylation of *HOXC10* and *HOXD1* showed the best performance in discriminating ESCC from normal samples by ROC curve analysis. Although hypermethylation events of the two genes were rarely reported in ESCC by previous studies, they have been extensively studied in other cancer types, especially in breast cancer [39,40]. Interestingly, in a recently published study, researchers have investigated the potential of *HOXC10* as a diagnostic marker for ESCC [41]. Using the WGBS technology, they identified the hypermethylated *HOXC10* in their cohort, which is consistent with the findings of this study.

Accumulating evidence demonstrated the crucial role of *HOXC10* in the development and progression of colorectal and gastric cancers [42,43]. *HOXC10* was reported upregulated in ESCC, and its high expression contributed to the proliferation and migration of tumor cells, indicating that *HOXC10* could be an unfavorable prognostic predictor [44]. The current findings revealed the pervasive hypermethylation status of *HOXC10* on ESCC, which did not seem to support the silenced expression of *HOXC10* by epigenetics regulation. Growing evidence has demonstrated the complex relationship between gene methylation and expression. The methylated CDKN2A gene (also known as p16 locus), which encoded two genes, p14ARF and p16INK4a, has been attempted as a screening marker for ESCC [45]. Interestingly, its promoter hypermethylation only silenced the expression of p14ARF but not p16INK4a [20], which implied that hypermethylation in different locations showed various effects on gene expression. In general, the promoter hypermethylation events can downregulate gene expression, but it is not always the case. For example, the hypermethylated *SDC2* has been successfully commercialized for the early detection of colorectal cancer [46-48], while an apparent contradiction is that its expression increased in colorectal cancer, and the upregulated expression promotes cancer development [49-51]. Relatively, the relationship between hypermethylation events in gene body and expression is more complex. It has been reported that the hypermethylation events in homeobox gene bodies did not suppress expression but rather upregulated the expression to activate their oncogene activity [52]. In this study, DMRs of *HOXC10* were primarily located in the gene body, which may explain this phenomenon. In addition, miRNA and lncRNA may involve in regulating gene expression too. In gastric cancer, *HOXC10* was found to be a direct target of MiR-136 [53]. The downregulated MiR-136 led to the upregulation of *HOXC10*, thus increasing the risk of peritoneal metastasis. The antisense transcript, lncHOXC-AS3, was also reported associated with the regulation of *HOXC10* expression [54] .

The hypermethylated *HOXD1* has been rarely reported on ESCC in previous studies. However, in colorectal cancer, its hypermethylation was associated with the silenced expression and occurred along with the cancer formation [55]. The hypermethylation was also observed in breast cancer and used as a biomarker to detect this disease [56,57]. In addition, methylated *HOXD1* was selected as a marker of lymph node metastasis in gastric cancer [58]. In this study, the Sanger sequencing results also revealed more frequently methylated events of *HOXD1* on ESCC samples than on normal samples. These findings suggested that *HOXD1* methylation might be a promising marker for ESCC detection.

We attempted to develop a classifier for ESCC by combing the methylation of *HOXL10* and *HOXD1*. The classifier obtained an AUC of 0.96 (95%CI: 0.91-0.99), with a sensitivity of 94.8% and specificity of 87.5% at the optimal threshold of 0.72 determined by the Youden index, suggesting its good performance in discriminating ESCC from normal samples. Furthermore, no significant variations were observed for the classifier in detecting ESCC with different age, gender, and pathological stages. However, the relationship between the classifier and patient features should be evaluated in a larger scale dataset as the results can be biased due to the small sample size. Notably, the classifier showed a lower detection rate for early-stage (I-II) than advanced stage ESCC (III-IV), which might be related to the higher methylation levels of *HOXL10* and *HOXD1* on advanced samples. Additionally, the classifier sensitivity for ESCA exceeded 80%, indicating its potential ability to detect ESCA.

Age is an important factor affecting DNA methylation. In the TCGA ESCC dataset (n = 94), we assessed the correlation of *HOXD1* and *GSX1* methylation with patient age. The *HOXD1* methylation showed a weak negative correlation with age (correlation coefficient = -0.19), but not significant (p = 0.056). While for *GSX1*, no correlation was observed (correlation coefficient = -0.055, p = 0.59), and this was the same for the other three genes, *HOXC11, HOXC10*, and *CDX21* (**supplementary figure 6**). The healthy controls included in this study were younger than ESCC patients because the healthy blood donors tended to be younger people. However, no significant correlation between methylation and age suggested that patient age may have limited effects on the methylation of these five genes.

Over 80% (n=40) of the HOXL subclass homeoboxes genes showed widespread hypermethylation in the nine most common cancer types, indicating their frequent methylation events in cancer cells. HOXL subclass genes belong to the superclass homeobox genes, which are characterized by sharing the homeobox sequence and play crucial roles in embryonic development and cell differentiation [59]. Many homeobox genes were found hypermethylated in different cancer types, including *HOXD1* and *HOXD10* [60,61] that were also identified in this study. In a pan-cancer study of more than 4000 genomic profiles [52], approximately 43% of homeobox genes were reported strong correlations between the overexpression and the gene body hypermethylation, suggesting DNA hypermethylation may be an epigenetic regulator of their upregulated expressions. Since many malignancy cells are tightly associated with stem cells, this can partially explain the frequent methylation events of homeobox genes in multiple cancer types [60].

## Conclusions

Genome-wide methylation profiling allowed us to interrogate the remodeling of DNA methylation during esophageal carcinogenesis from a landscape view. We observed the widespread hypomethylation events and frequent hypermethylation of HOXL subclass homeoboxes and Zinc finger family genes in ESCC. Two HOXL subclass homeoboxes, *HOXC10* and *HOXD1*, presented good classification abilities for ESCC and normal samples. Further investigations indicated that frequent hypermethylation of HOXL subclass homeoboxes occurred in multiple cancer types, and one of its members, *GSX1*, whose methylation status was verified by sequencing, could be a maker candidate for pan-cancer detection. Early detection of ESCC is a challenging task. Many previous studies in epigenetics have paved a concrete road for unraveling the mechanism of ESCC carcinogenesis and identifying high-performed diagnostic markers. Our findings provide new insights to understand the epistatic remodeling and discover new methylation biomarkers for ESCC.

## Data Availability

The TCGA CRC 450k data and GEO datasets are publicly available online. The samples and data generated in this study have been fully reflected in the manuscript.

## List of abbreviations

EC: esophageal cancer
ESCA: esophageal adenocarcinoma
ESCC: esophageal squamous cell carcinoma
DMC: differentially methylated CpGs
DMR: differentially methylated regions
DMG: differentially methylated genes
WGBS: whole-genome bisulfite sequencing
TCGA: The Cancer Genome Atlas
MSP: methylation specific PCR

## Declarations

### Ethics approval and consent to participate

Approval for this study was obtained from the Ethics Committee of the First Affiliated Hospital of Zhengzhou University (approval number 2020-KY-0152). All experiments were performed in accordance with relevant guidelines and regulations. Written informed consent was obtained from individual or guardian participants.

### Consent for publication

The current manuscript has been read and approved by all named authors.

### Authors’ contributions

XP L conceived and supervised the study; QN Y and NM X designed experiments; YT Z and HF J analyzed data; RY C, FL Y, LHY C and Y X performed experiments; J Z and DH Z provided new tools and reagents; QN Y, NM X and KK W wrote the manuscript; XP L made manuscript revisions.

