## supplemental figures and tables for "Genome-wide methylation profiling identify hypermethylated HOXL subclass genes as potential markers for esophageal squamous cell carcinoma detection"

### Supplementary figures

**Supplementary figure 1. DMCs genomic distribution characteristics.**

**A:** The distribution of NC-DMCs and ESCC-DMCs on different chromosomes. **B:** Distance of adjacent ESCC-DMCs and adjacent NC-DMCs across different chromosomes. **C:** The correlation coefficients of adjacent ESCC-DMCs and NC-DMCs.

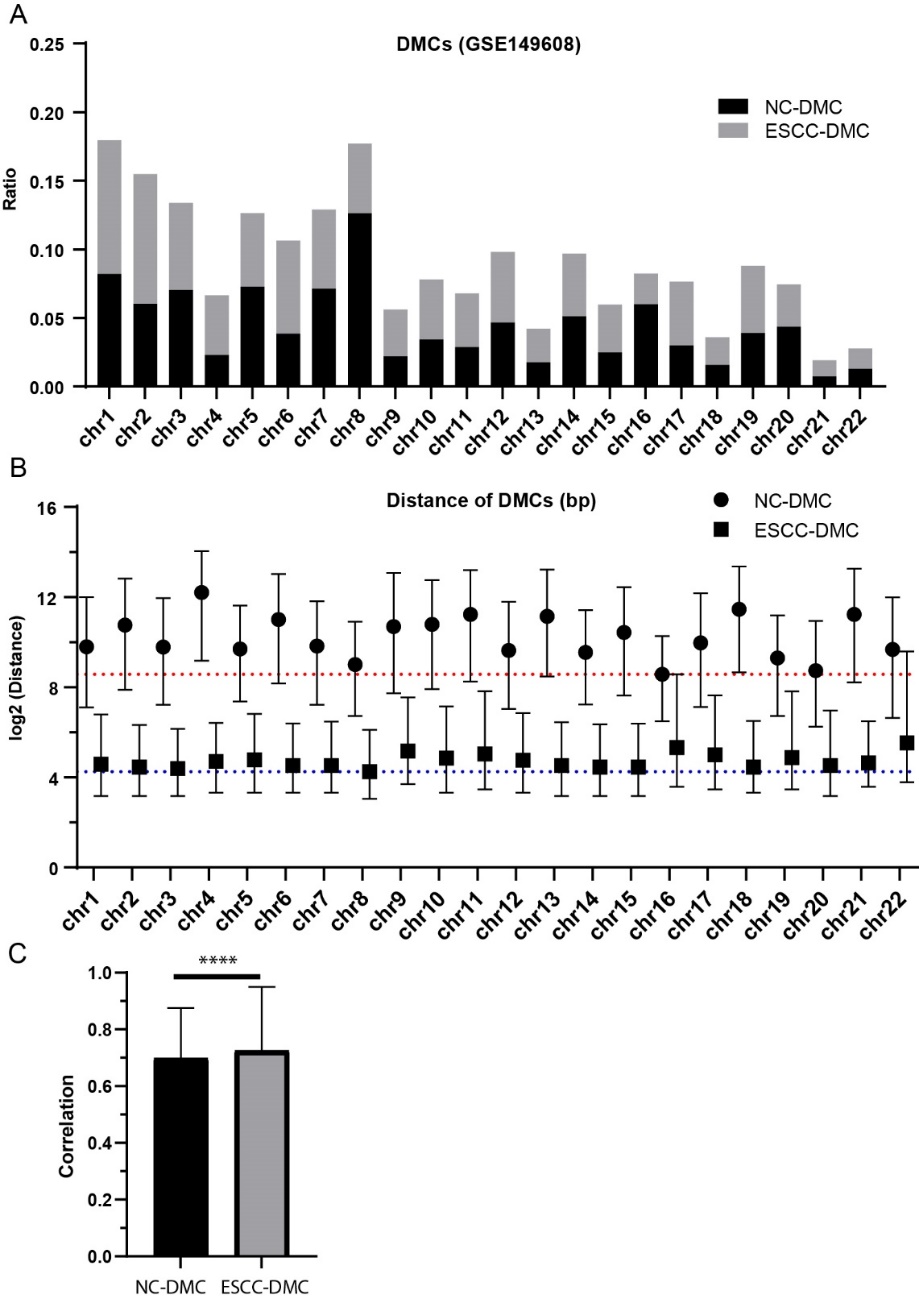

**Supplementary figure 2. DMRs and DMGs genomic distribution characteristics.**

**A:** The distribution of NC-DMRs and ESCC-DMRs on different chromosomes. **B:** Distance of adjacent ESCC-DMRs and adjacent NC-DMRs across different chromosomes. **C:** The frequency of NC-DMGs and ESCC-DMGs across different chromosomes.

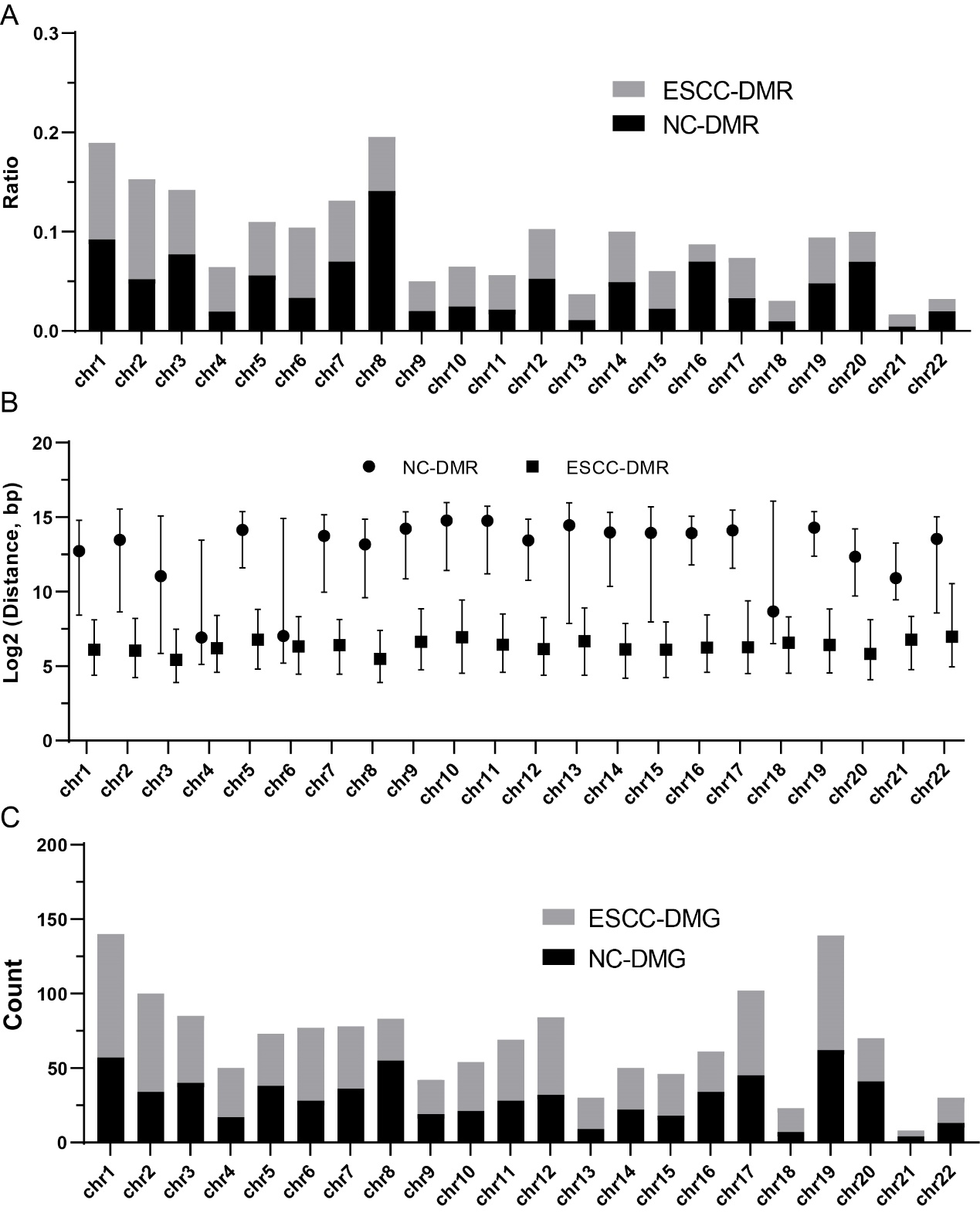

**Supplementary figure 3. Similarity of the normal and ESCC samples in GSE52826 dataset. A:** tree plot showing the hierarchical clustering results. **B:** t-SNE analysis showing the difference of normal and ESCC samples.

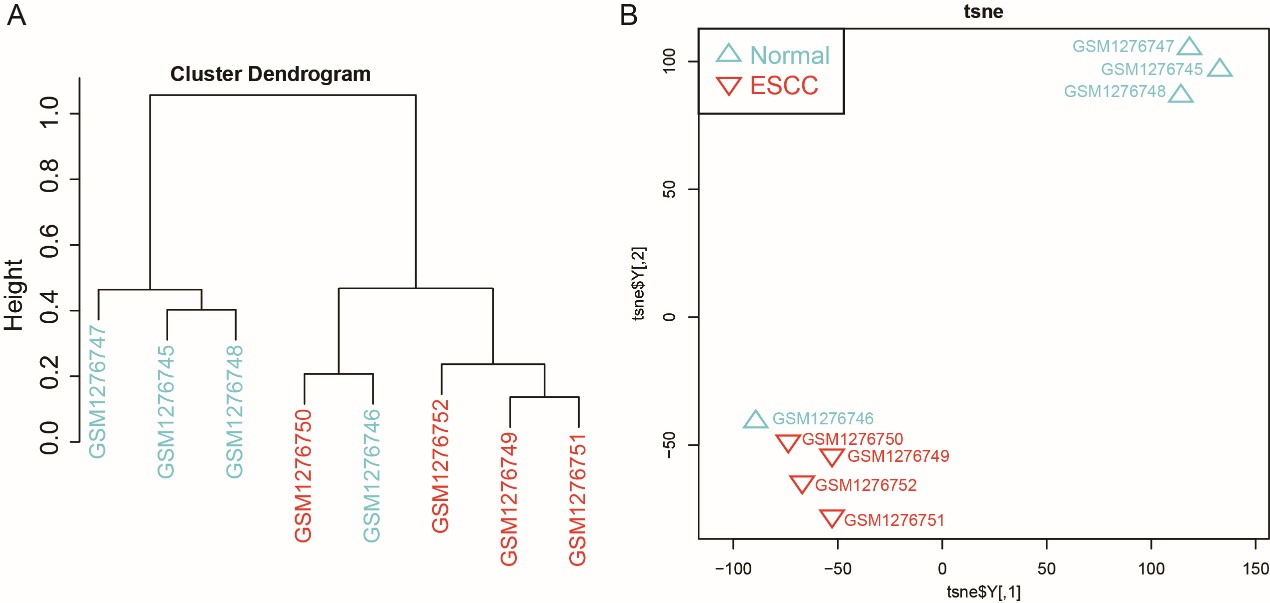

**Supplementary figure 4. The methylation levels of *HOXC10* (A) and *HOXD1* (B) in different stages of ESCC.** Kruskal test was used for multiple group comparisons.

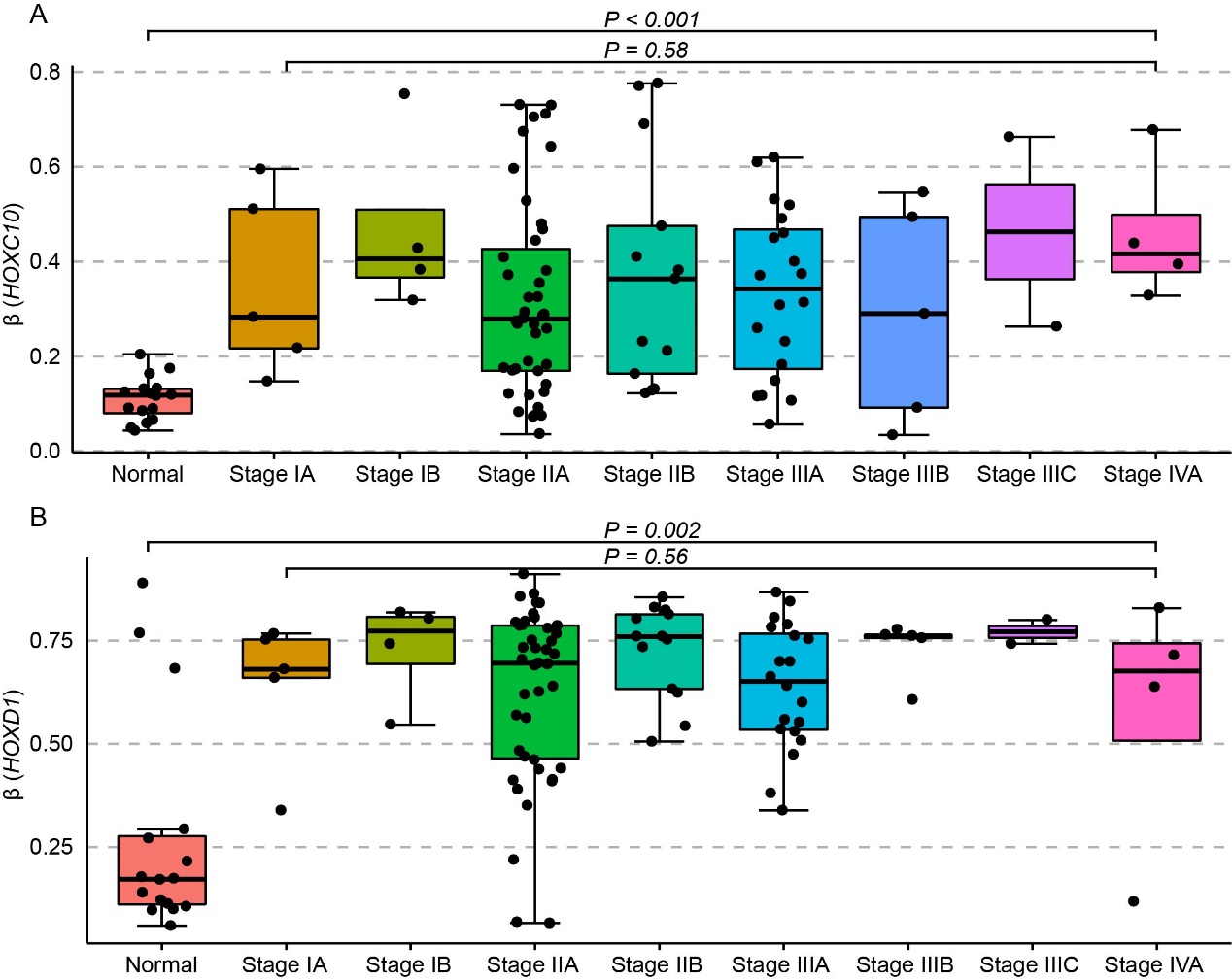

**Supplementary figure 5. ROC curves of the 13 potential methylation markers showed the diagnostic performance for ESCC.** **A-M:** *CDKN2A*, *CDKN2B*, *TFF1*, *MGMT*, *MLH1*, *DAPK1*, *SCGB3A1*, *TFPI2*, *DACH1*, *SOX17*, *CHFR*, *CDH1*, *APC*. The probe with the most significant differences between ESCC and normal were selected for each gene. AUC values and the 95% confidence interval (CI) were also calculated.

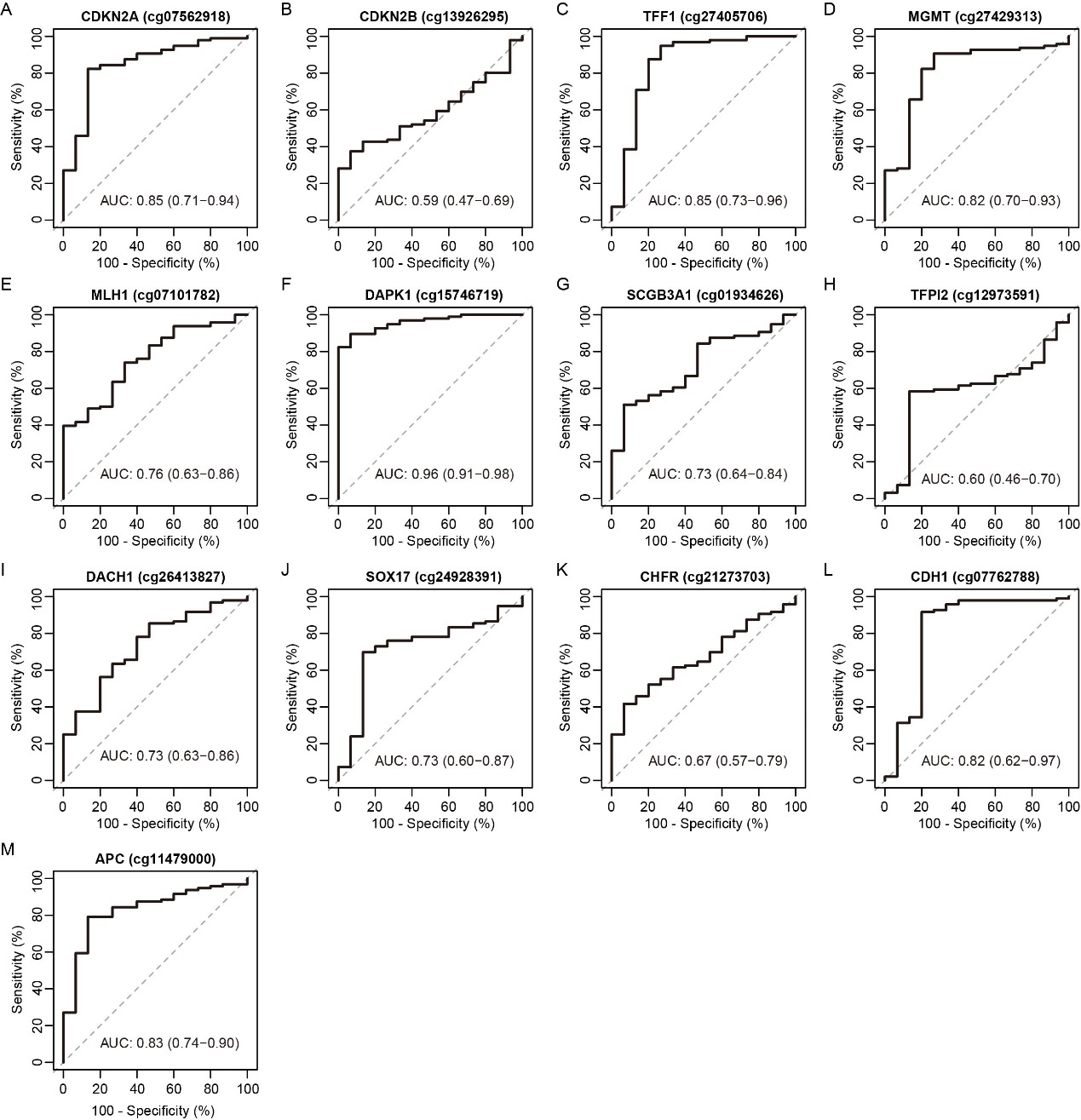

**Supplementary figure 6. The correlation of methylation level with patient age.** Pearson’s correlation coefficient was estimated for each gene.

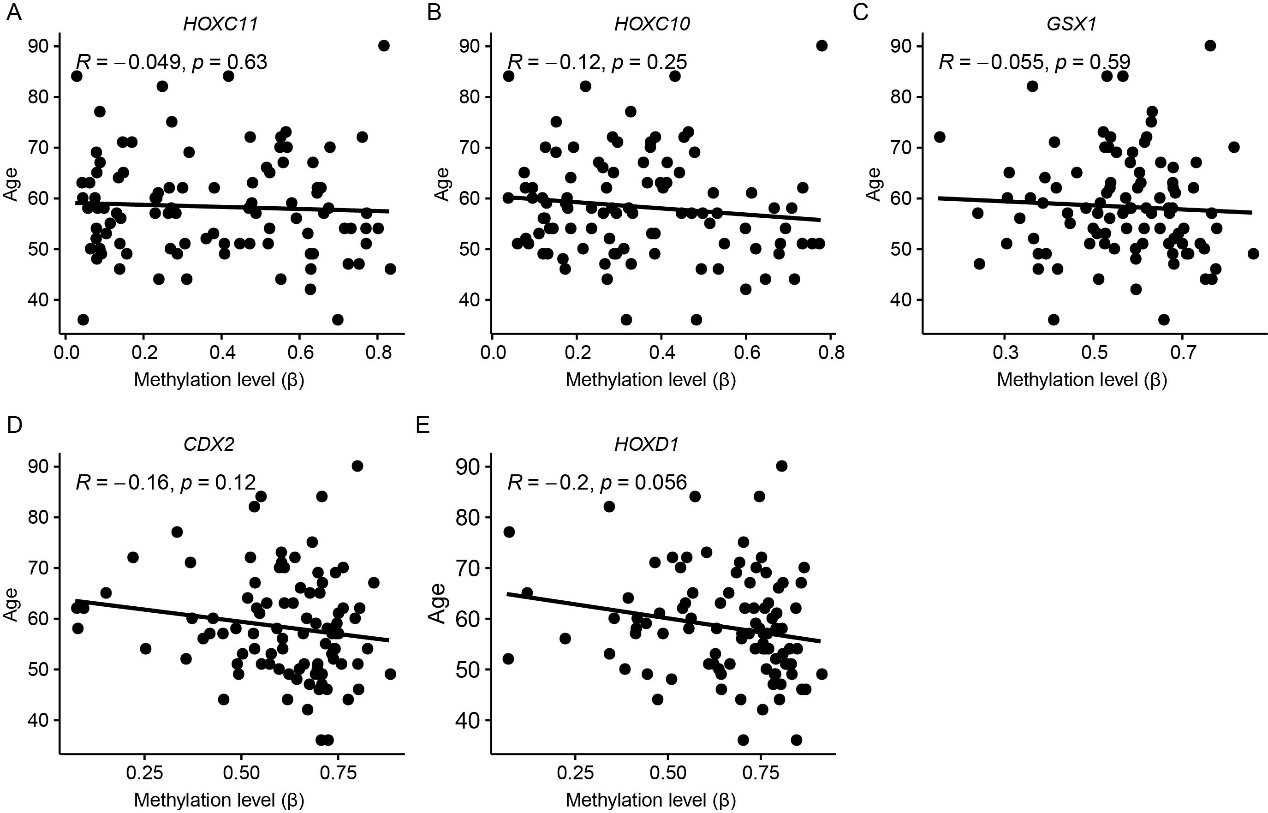

### Supplementary tables

**Supplementary table 1. Clinical features of GSE149608 dataset.**

| Specimen | Age (year, range) | Sex | TNM stage | Lymph node metastasis | Histologic stage |
| --- | --- | --- | --- | --- | --- |
| P1 | 61-65 | male | T3N1aM0 | Yes | G2 |
| P2 | 76-80 | female | T3N0M0 | No | G3 |
| P3 | 56-60 | male | T2N0M0 | No | G2 |
| P4 | 56-60 | female | T3N1aM0 | Yes | G2 |
| P5 | 56-60 | male | T3N0M0 | No | G2 |
| P6 | 61-65 | male | T3N0M0 | No | G1 |
| P7 | 55-60 | male | T3N0M0 | No | G2 |
| P8 | 61-65 | male | T2N1aM0 | Yes | G3 |
| P10 | 56-60 | female | T1bN0M0 | Yes | G2 |

**Supplementary table 2. Clinical features of GSE52826 dataset.**

| Specimen | Age (year, range) | Sex | TNM stage | Histologic grade | Tumor location | Stage |
| --- | --- | --- | --- | --- | --- | --- |
| T1 | 61-65 | male | T2N0M0 | G1 | Lower | IB |
| T2 | 61-65 | male | T1N0M0 | G2 | Middle | IB |
| T3 | 61-65 | female | T1N0M0 | G1 | Middle | IA |
| T4 | 61-65 | male | T2N0M0 | G1 | Middle | IIA |

**Supplementary table 3. Clinical features of the 33 ESCC samples and 20 healthy blood samples.**

| Sample ID | Age (year, range) | Sex | Grade | TNM Stage | Lymph node metastases | Sample type |
| --- | --- | --- | --- | --- | --- | --- |
| BN01 | 31-35 | Female |  |  |  | Healthy |
| BN02 | 21-25 | Male |  |  |  | Healthy |
| BN03 | 26-30 | Male |  |  |  | Healthy |
| BN04 | 36-40 | Male |  |  |  | Healthy |
| BN05 | 21-25 | Male |  |  |  | Healthy |
| BN06 | 31-35 | Male |  |  |  | Healthy |
| BN07 | 36-40 | Female |  |  |  | Healthy |
| BN08 | 31-35 | Male |  |  |  | Healthy |
| BN09 | 21-25 | Male |  |  |  | Healthy |
| BN10 | 31-35 | Female |  |  |  | Healthy |
| BN11 | 36-40 | Male |  |  |  | Healthy |
| BN12 | 31-35 | Male |  |  |  | Healthy |
| BN13 | 41-45 | Male |  |  |  | Healthy |
| BN14 | 26-30 | Male |  |  |  | Healthy |
| BN15 | 26-30 | Male |  |  |  | Healthy |
| BN16 | 36-40 | Male |  |  |  | Healthy |
| BN17 | 26-30 | Male |  |  |  | Healthy |
| BN18 | 26-30 | Male |  |  |  | Healthy |
| BN19 | 26-30 | Male |  |  |  | Healthy |
| BN20 | 26-30 | Male |  |  |  | Healthy |
| EN1 |  |  |  |  |  | Adjacent |
| EN2 |  |  |  |  |  | Adjacent |
| EN3 |  |  |  |  |  | Adjacent |
| EN4 |  |  |  |  |  | Adjacent |
| EN5 |  |  |  |  |  | Adjacent |
| EN6 |  |  |  |  |  | Adjacent |
| EN7 |  |  |  |  |  | Adjacent |
| EN8 |  |  |  |  |  | Adjacent |
| EN9 |  |  |  |  |  | Adjacent |
| EN10 |  |  |  |  |  | Adjacent |
| EN11 |  |  |  |  |  | Adjacent |
| EN12 |  |  |  |  |  | Adjacent |
| EN13 |  |  |  |  |  | Adjacent |
| EN14 |  |  |  |  |  | Adjacent |
| EN15 |  |  |  |  |  | Adjacent |
| EN16 |  |  |  |  |  | Adjacent |
| EN17 |  |  |  |  |  | Adjacent |
| EN18 |  |  |  |  |  | Adjacent |
| EN19 |  |  |  |  |  | Adjacent |
| EN20 |  |  |  |  |  | Adjacent |
| ESCC1 | 51-55 | Male | G1 | pT3N0Mx | NO | ESCC tissue |
| ESCC2 | 56-60 | Male | G2 | pT3N2Mx | YES | ESCC tissue |
| ESCC3 | 61-65 | Male | G2 | na | YES | ESCC tissue |
| ESCC4 | 61-65 | Male | G2 | PT3N3Mx | YES | ESCC tissue |
| ESCC5 | 66-70 | Male | G2 | pT2N1Mx | YES | ESCC tissue |
| ESCC6 | 66-70 | Male | G2 | na | NO | ESCC tissue |
| ESCC7 | 61-65 | Male | G1 | pT3N1Mx | YES | ESCC tissue |
| ESCC8 | 46-50 | Male | G2 | pT1bN0Mx | NO | ESCC tissue |
| ESCC9 | 71-75 | Male | G2 | pT2N0Mx | NO | ESCC tissue |
| ESCC10 | 61-65 | Male | G1 | pT3N0Mx | NO | ESCC tissue |
| ESCC11 | 66-70 | Male | G1 | pT3N1Mx | YES | ESCC tissue |
| ESCC12 | 56-60 | Male | G3 | pT3N1Mx | YES | ESCC tissue |
| ESCC13 | 66-70 | Male | G2 | pT3N1Mx | YES | ESCC tissue |
| ESCC14 | 71-75 | Female | G1 | pT1bNoMx | NO | ESCC tissue |
| ESCC15 | 56-60 | Female | G2 | pT1aN0Mx | NO | ESCC tissue |
| ESCC16 | 66-70 | Male | G2 | pT3N1Mx | YES | ESCC tissue |
| ESCC17 | 61-65 | Female | G2 | pT3N1Mx | YES | ESCC tissue |
| ESCC18 | 61-65 | Male | G3 | na | YES | ESCC tissue |
| ESCC19 | 61-65 | Male | G2 | pT3N0Mx | NO | ESCC tissue |
| ESCC20 | 51-55 | Male | G1 | pT3N3Mx | YES | ESCC tissue |
| ESCC21 | 51-55 | Male | G2 | pT3N0Mx | NO | cfDNA |
| ESCC22 | 66-70 | Male | NA | pT2N1Mx | YES | cfDNA |
| ESCC23 | 66-70 | Female | G1 | pT2N0Mx | NO | cfDNA |
| ESCC24 | 66-70 | Female | G2 | pT3N1Mx | YES | cfDNA |
| ESCC25 | 51-55 | Male | G2 | pT3N3Mx | YES | cfDNA |
| ESCC26 | 66-50 | Female | G2 | pT3N0Mx | NO | cfDNA |
| ESCC27 | 41-45 | Male | G1 | pT2N2Mx | YES | cfDNA |
| ESCC28 | 56-60 | Male | G1 | pT3N1Mx | YES | cfDNA |
| ESCC29 | 71-75 | Female | G1 | pT3N0Mx | NO | cfDNA |
| ESCC30 | 51-55 | Male | G1 | pT3N2Mx | YES | cfDNA |
| ESCC31 | 56-60 | Male | G2 | pT3N0Mx | NO | cfDNA |
| ESCC32 | 61-65 | Male | G2 | pT1bN0Mx | NO | cfDNA |
| ESCC33 | 56-60 | Female | G1 | pT3N0Mx | NO | cfDNA |

**Supplementary table 4. The MSP amplification regions of *HOXD1* and *GSX1*.**

| DMR_id | Postion | Methy (N, %) | Methy (ESCC, %) | CpG_count | delta (Methy) |
| --- | --- | --- | --- | --- | --- |
| HOXD1-R1 | chr2: 177054188-177054237 | 3.73 | 27.45 | 3 | 23.72 |
| HOXD1-R2 | chr2: 177054245-177054282 | 9.51 | 35.52 | 4 | 26 |
| HOXD1-R3 | chr2: 177054303-177054350 | 5.85 | 27.72 | 5 | 21.87 |
| HOXD1-R4 | chr2: 177054495-177054547 | 13.09 | 47.81 | 5 | 34.72 |
| HOXD1-R5 | chr2: 177054572-177054585 | 14.53 | 47.66 | 3 | 33.13 |
| HOXD1-R6 | chr2: 177054648-177054696 | 11.05 | 40.36 | 5 | 29.31 |
| HOXD1-R7 | chr2: 177054836-177054888 | 19.73 | 46.52 | 3 | 26.78 |
| GSX1-DMR16 | chr13: 28367740-28367789 | 13.54 | 37.35 | 5 | 23.8 |
| GSX1-DMR17 | chr13: 28367815-28367852 | 10.37 | 34.46 | 5 | 24.09 |

**Supplementary table 5. Summary of DMCs and DMRs in different genomic regions.**

|  | NC-DMC | ESCC-DMC | NC-DMR | ESCC-DMR |
| --- | --- | --- | --- | --- |
| Upstream (0-2kb) | 72592 | 14254 | 1294 | 1819 |
| innergenic | 88544 | 30002 | 1734 | 4018 |
| Downstream (200bp) | 74686 | 11069 | 1323 | 1396 |
| intergenic | 125519 | 13953 | 2071 | 1807 |

**Supplementary table 6. The identified NC-DMGs and ESCC-DMGs in this study.**

|  | Genes |
| --- | --- |
| NC-DMGs | MXRA8, AL645728.1, SLC35E2B, TTC34, CHD5, TNFRSF9, CA6, SPSB1, TNFRSF8, TNFRSF1B, AADACL4, FHAD1, ZBTB17, MRTO4, MINOS1-NBL1, CDA, RAP1GAP, EPHA8, HPCA, C1orf94, ZSWIM5, TTC39A, IFI44, SYDE2, RP11-302M6.4, NTNG1, KCNA2, S100A13, RAB13, KCNN3, MSTO1, PEAR1, ATP1A4, F11R, MGST3, FAM78B, DPT, DNM3, TNFSF4, TNR, CFHR3, NR5A2, PKP1, SYT2, TMCC2, LEMD1, SPATA17, LYPLAL1, C1orf65, GGPS1, FMN2, RGS7, SCCPDH, NLRP3, GCSAML, OR2G6, LYPD8, FAM110C, MYT1L, FAM84A, DRC1, KCNK3, SLC35F6, ALK, LTBP1, PSME4, SPTBN1, LGALSL, SPRED2, RTKN, HTRA2, TRABD2A, CAPG, KRCC1, SMYD1, KIAA1211L, IL1R1, FHL2, UXS1, PSD4, CBWD2, NCKAP5, CACNB4, DAPL1, LRP2, SP9, HECW2, PLCL1, WNT6, ANKMY1, BOK, TRANK1, SNRK, PDZRN3, CRYBG3, IMPG2, CEP97, DPPA2, SPICE1, ADCY5, SLC12A8, ALG1L, CHCHD6, DNAJB8, RPN1, EFCC1, COL6A5, NEK11, MRPS22, PCOLCE2, AGTR1, CP, MME, IQCJ-SCHIP1, NLGN1, KCNMB3, ZNF639, PEX5L, MCF2L2, ABCC5, IL1RAP, GMNC, MB21D2, HES1, TNK2, SLC51A, PCYT1A, NRROS, PIGX, NCBP2, MFI2, RNF212, SPON2, JAKMIP1, AFAP1, LDB2, FAM184B, SEL1L3, GRXCR1, FRYL, FIP1L1, NOA1, IGFBP7, LPHN3, MOB1B, MAPK10, HSPA4L, PDGFC, SLC6A19, SLC6A18, TERT, MRPL36, UBE2QL1, ADCY2, ROPN1L, RP11-1C1.5, ANKRD33B, DNAH5, FBXL7, 11-Mar, CDH12, CDH6, PDZD2, NPR3, ADAMTS12, AGXT2, FYB, PLCXD3, HCN1, CTD-2117L12.1, FOXD1, ATP6AP1L, NUDT12, SNCAIP, ZNF608, CDC42SE2, KCTD16, JAKMIP2, HTR4, ABLIM3, ADRA1B, GABRB2, RANBP17, MSX2, UNC5A, RASGEF1C, DUSP22, NQO2, CDYL, F13A1, TFAP2A, NEDD9, JARID2, NRSN1, ALDH5A1, HIST1H2BC, PPARD, C6orf89, DAAM2, TREML2, GNMT, PTK7, TMEM151B, RCAN2, FBXO9, COL19A1, PGM3, KIAA1009, GJB7, MAN1A1, PKIB, TPD52L1, SMOC2, WDR27, PRKAR1B, ADAP1, AMZ1, FOXK1, MMD2, RNF216, DAGLB, CCDC129, PDE1C, AVL9, BMPER, POU6F2, HECW1, PSPH, ZNF479, ZNF716, KCTD7, WBSCR17, SRRM3, PPP1R9A, SHFM1, BAIAP2L1, MUC17, PRKRIP1, RELN, LHFPL3, MET, SPAM1, GRM8, PLXNA4, ATP6V0A4, DPP6, PAXIP1-AS2, BLACE, DNAJB6, PTPRN2, DLGAP2, CSMD1, MTMR7, DPYSL2, EXTL3, CHRNB3, SNAI2, SNTG1, ST18, FAM150A, XKR4, FAM110B, CLVS1, NKAIN3, DNAJC5B, ADHFE1, COPS5, CPA6, C8orf34, XKR9, KCNB2, GDAP1, MRPS28, CA3, CNBD1, MMP16, CPQ, C8orf47, NIPAL2, SPAG1, NCALD, RSPO2, PKHD1L1, CSMD3, SAMD12, COL14A1, ZHX2, FAM83A, ANXA13, TRMT12, OC90, KCNQ3, LRRC6, KHDRBS3, FAM135B, COL22A1, KCNK9, TRAPPC9, DENND3, AC138647.1, TSNARE1, ARC, GML, FAM83H, KIFC2, LINGO2, RP11-613M10.9, ROR2, FGD3, PTPDC1, ZFP37, ASTN2, BRINP1, PSMB7, FAM102A, NAIF1, RP11-101E3.5, C9orf171, GFI1B, RXRA, LCN1, KCNT1, GRIN1, PNPLA7, CUBN, ST8SIA6, SLC39A12, HK1, UNC5B, PLA2G12B, NRG3, CRTAC1, PAX2, INA, SH3PXD2A, ITPRIP, SORCS1, ATRNL1, PNLIPRP3, DOCK1, PTPRE, TTC40, GPR123, KNDC1, SPRN, MUC2, MUC5B, TOLLIP, BRSK2, KCNQ1, PHLDA2, CARS, ARNTL, TSPAN18, PRDM11, SLC39A13, PRG3, MS4A6E, SLC22A11, SYT12, C11orf72, FGF3, ANO1, PDE2A, POLD3, SLCO2B1, LRRC32, PIWIL4, COLCA1, NCAM1, ZBTB16, DSCAML1, KCNJ1, CACNA2D4, CACNA1C, RP11-234B24.6, ANO2, PPFIBP1, CPNE8, PRR13, DNAJC14, NDUFA4L2, TBC1D30, GRIP1, CPM, PLXNC1, NR1H4, C12orf42, WSCD2, CMKLR1, SELPLG, C12orf76, CUX2, TBX3, CCDC60, P2RX7, HPD, HCAR1, TMEM132B, TMEM132C, GLT1D1, TMEM132D, FZD10, GPR133, GALNT9, ALOX5AP, KBTBD6, KBTBD7, WDFY2, CLN5, KDELC1, FAM155A, MYO16, TMEM255B, SLC7A8, AKAP6, NPAS3, SLC25A21, MDGA2, RGS6, ACOT6, NRXN3, GALC, PSMC1, SLC24A4, RIN3, ITPK1, RP11-371E8.4, UNC79, TCL1A, WDR25, BEGAIN, DLK1, CDC42BPB, KLC1, TMEM179, GOLGA8J, OTUD7A, CHRNA7, C15orf52, BAHD1, PARP16, ANP32A, HCN4, ISLR2, CCDC33, CHRNB4, HDGFRP3, SLC28A1, AGBL1, NTRK3, SV2B, NR2F2, ADAMTS17, RHBDF1, CACNA1H, GNPTG, MEFV, ADCY9, ALG1, RBFOX1, GRIN2A, XYLT1, UMOD, DNAH3, NDUFAB1, DCTN5, PRKCB, CACNG3, AQP8, HS3ST4, IL21R, GSG1L, C16orf92, SEPHS2, ZNF689, PRSS8, TGFB1I1, ZNF423, CDH11, WDR59, RP11-77K12.1, OSGIN1, KIAA0513, JPH3, CBFA2T3, CDH15, PRDM7, RAP1GAP2, GLTPD2, AIPL1, PIK3R5, USP43, RCVRN, GAS7, MYOCD, SLC5A10, MAP2K3, PIPOX, SSH2, ASIC2, CDC6, TNS4, KRT17, AOC2, WNT3, WNT9B, ITGB3, GIP, SGCA, CA10, ANKFN1, EPX, PPM1E, ABCA10, CD300E, RAB37, GRIN2C, NUP85, ZACN, MXRA7, MGAT5B, TNRC6C, TMC8, C17orf99, AFMID, RBFOX3, CARD14, AATK, TMEM105, FSCN2, ASPSCR1, TBCD, EMILIN2, PIEZO2, HRH4, CCDC178, RIT2, SLC14A2, DCC, SHC2, ODF3L2, TPGS1, PRTN3, CELF5, SEMA6B, CATSPERD, TNFSF14, CTB-133G6.1, PEX11G, C19orf45, EVI5L, FBN3, CACNA1A, EMR3, CPAMD8, NR2F6, PLVAP, SLC27A1, UNC13A, GDF15, COPE, MEF2BNB-MEF2B, HAPLN4, CHST8, PRODH2, RYR1, CTC-435M10.3, CEACAM4, CD79A, LYPD3, CKM, NOVA2, C5AR1, SLC8A2, BSPH1, ELSPBP1, SYT3, GPR32, CD33, SIGLEC10, SIGLEC5, FPR3, PRKCG, VSTM1, LILRB5, LILRB2, LILRA4, LAIR1, LAIR2, KIR3DX1, LILRA2, LILRB1, KIR3DL1, NLRP7, TMEM150B, NLRP4, NLRP8, SMIM17, ZIM2, ZSCAN1, SLC27A5, TMC2, PAK7, OVOL2, THBD, DEFB121, SUN5, BPIFB6, BPIFA2, BPIFA3, AHCY, MYH7B, SCAND1, DLGAP4, VSTM2L, KIAA1755, BPI, ARHGAP40, MAFB, PTPRT, WFDC10B, PREX1, PTGIS, FAM65C, DOK5, CASS4, GNAS, ZNF831, PHACTR3, CDH26, CDH4, HRH3, C20orf166, NTSR1, NKAIN4, COL20A1, KCNQ2, EEF1A2, PTK6, GMEB2, ZBTB46, MYT1, LIPI, C21orf49, RIPK4, ITGB2, GAB4, CECR2, RTN4R, SGSM1, SEZ6L, AP1B1, TXN2, IL2RB, CACNA1I, KIAA0930, C22orf34, TTLL8, MOV10L1, DMD, MAMLD1, |
| ESCC-DMGs | PLEKHN1, AGRN, TMEM240, PLCH2, TNFRSF14, WRAP73, AJAP1, ACOT7, ESPN, PLEKHG5, KLHL21, KAZN, HSPB7, CROCC, PAX7, CAPZB, HTR6, VWA5B1, CAMK2N1, ALPL, LDLRAD2, LACTBL1, STPG1, RUNX3, MAP3K6, WASF2, CCDC28B, SMIM12, RSPO1, KCNQ4, C1orf50, PTPRF, BEST4, PTCH2, POMGNT1, FOXE3, FOXD2, DMRTA2, ZYG11A, TTC22, LEPR, C1orf141, ST6GALNAC5, PTGFR, GFI1, GPR88, STRIP1, KCNA3, ADORA3, SYT6, ANKRD35, C2CD4D, TCHH, NPR1, PKLR, FDPS, SEMA4A, HAPLN2, BCAN, LRRC71, KLHDC9, CCDC181, LHX4, KIAA1614, CACNA1E, ZNF648, RGS2, CFHR2, GPR25, SHISA4, BTG2, KLHDC8A, RASSF5, SERTAD4, KCNK2, ESRRG, HLX, CAPN2, ITPKB, WNT3A, GUK1, ACTA1, TRIM58, SOX11, OSR1, GDF7, KLHL29, UCN, CYP1B1, ARHGEF33, TMEM178A, SIX3, SIX2, NRXN1, EFEMP1, OTX1, MEIS1, CNRIP1, VAX2, AC007040.11, CYP26B1, EMX1, NOTO, EGR4, C2orf81, LBX2, TLX2, LOXL3, CTNNA2, VAMP8, VAMP5, CD8A, ITPRIPL1, ANKRD23, AC092675.3, NCK2, PAX8, EN1, SCTR, GLI2, GYPC, PTPN18, ARHGEF4, CCDC74A, NXPH2, DLX2, EVX2, HOXD13, HOXD12, HOXD10, HOXD3, HOXD1, ZNF804A, TMEFF2, BOLL, C2orf47, TNS1, CYP27A1, WNT10A, DES, PAX3, CCDC140, ECEL1, GBX2, ACKR3, MLPH, LRRFIP1, RAMP1, AC016757.3, CLASP2, CCK, PTH1R, MST1, RNF123, ARHGEF3, FEZF2, PRICKLE2, BBX, DPPA4, BOC, LSAMP, CASR, PDIA5, ROPN1B, KLF15, PLXNA1, ABTB1, KBTBD12, RUVBL1, GATA2, TRH, SOX14, ESYT3, FOXL2, C3orf72, SPSB4, GRK7, CHST2, PLSCR2, ZIC4, ZIC1, GPR149, SHOX2, GFM1, C3orf80, SLITRK3, LRRC34, GHSR, SST, LPP, FGF12, GP5, FAM43A, XXYLT1, MFSD7, RGS12, ADRA2C, MSX1, HMX1, SLC2A9, NKX3-2, FAM200B, PROM1, SOD3, PHOX2B, BEND4, SHISA3, SLC10A4, NPFFR2, CXCL5, CXCL2, FGF5, NKX6-1, BANK1, TACR3, DKK2, TRAM1L1, NDNF, TNIP3, TMEM155, TRPC3, PCDH10, MGARP, RP11-542P2.1, UCP1, POU4F2, HAND2, IRX4, C5orf38, RXFP3, PTGER4, MCIDAS, CCNO, OTP, ANKRD34B, NR2F1, GPR150, KCNN2, CDO1, SHROOM1, CTC-349C3.1, H2AFY, NEUROG1, LECT2, PROB1, CXXC5, NRG2, PCDHA1, PCDHGA1, RBM27, CD74, MYOZ3, SPARC, HAND1, SOX30, C5orf52, TLX3, NKX2-5, DRD1, HRH2, GPRIN1, MAML1, MGAT4B, IRF4, FOXF2, TUBB2B, PXDC1, PPP1R3G, NRN1, RREB1, SYCP2L, DCDC2, HIST1H4F, BTN1A1, HIST1H2BK, POM121L2, ZSCAN31, TULP1, SRSF3, FOXP4, TFEB, GUCA1A, TTBK1, NFKBIE, TFAP2D, DST, COL9A1, COL12A1, HTR1B, TBX18, CNR1, EPHA7, PRDM13, MCHR2, BVES, NR2E1, GPR6, DSE, FAM162B, VGLL2, CLVS2, RSPO3, SOGA3, TMEM200A, TCF21, MAP3K5, SLC35D3, OLIG3, NMBR, ULBP1, T, THBS2, FBXL18, GRID2IP, TWIST1, TMEM196, SP8, NPY, HOXA2, HOXA9, HOXA13, EVX1, PRR15, CRHR2, YAE1D1, CAMK2B, SUN3, C7orf57, VSTM2A, EGFR, ZNF273, AC104057.1, LIMK1, CLIP2, SEMA3D, GRM3, ABCB1, GNGT1, PDK4, DLX5, TAC1, MBLAC1, AGFG2, FEZF1, TMEM229A, PRRT4, FLNC, FAM115A, RARRES2, ZNF775, NOS3, EN2, SHH, VIPR2, SOX7, PHYHIP, NKX2-6, PNMA2, ZNF395, PURG, GPR124, ADRB3, SOX17, MOS, PENK, CRH, RP11-1102P16.1, RP11-383H13.1, STMN2, ATP6V0D2, GDF6, TSPYL5, STK3, OSR2, C8orf56, TRPS1, AARD, MAFA, MROH6, SCRT1, FOXH1, LRRC14, FOXD4, NFIB, PLIN2, ACER2, KLF9, FOXB2, C9orf129, BARX1, TRAF1, DAB2IP, LHX6, LHX2, NR5A1, OLFML2A, LMX1B, DNM1, BARHL1, RALGDS, ADAMTS13, LHX3, TRAF2, TOR4A, NSMF, KLF6, C1QL3, GAD2, FZD8, RET, HNRNPF, ZNF239, GDF10, CHAT, PHYHIPL, TMEM26, NEUROG3, ZMIZ1, PLAC9, CDHR1, CH25H, CYP26C1, CYP26A1, NKX2-3, ABCC2, SEC31B, TLX1, LBX1, FGF8, TRIM8, VAX1, EMX2, TACC2, HMX3, HMX2, EBF3, TCERG1L, PWWP2B, NLRP6, RNH1, C11orf35, MOB2, DUSP8, SYT8, IGF2, CD81, PRKCDBP, AMPD3, CALCB, MYOD1, SLC6A5, NELL1, SLC17A6, BDNF, KCNA4, PAX6, RCN1, WT1, ABTB2, RAG1, ALX4, CREB3L1, DGKZ, LRRC10B, FADS2, AHNAK, HRASLS5, MACROD1, C11orf85, TIGD3, CFL1, ADRBK1, TCIRG1, SHANK2, CCDC67, CRYAB, BSX, ROBO3, ST3GAL4, RAD52, KCNA5, VWF, RERG, BCAT1, SLC38A1, SLC38A4, HDAC7, DKFZP779L1853, WNT1, DDN, C1QL4, KCNH3, RACGAP1, METTL7A, GALNT6, GRASP, KRT86, RARG, AAAS, ATP5G2, HOXC13, HOXC12, HOXC11, HOXC10, RP11-834C11.12, HOXC4, B4GALNT1, CTDSP2, AVPR1A, HMGA2, TRHDE, MYF6, MYF5, ACSS3, SLC6A15, ALX1, USP44, PAH, ASCL1, TXNRD1, CHST11, RP11-144F15.1, ASCL4, FOXN4, FAM222A, SH2B3, LHX5, MSI1, KDM2B, RHOF, CHFR, TNFRSF19, AMER2, GSX1, PDX1, CDX2, URAD, FLT1, SERTM1, SERP2, KIAA0226L, MLNR, LECT1, PCDH8, PCDH17, KLHL1, EDNRB, SLITRK1, SLITRK5, CLDN10, FGF14, RAB20, SLC7A7, JPH4, REC8, RIPK3, FOXG1, C14orf23, NFKBIA, INSM2, PAX9, FOXA1, SSTR1, CLEC14A, PTGDR, PTGER2, OTX2, L3HYPDH, C14orf39, SIX1, SLC8A3, VSX2, GPR68, GSC, BCL11B, SLC25A29, AMN, EXOC3L4, TEX22, CRIP1, GABRB3, GOLGA8M, KLF13, GOLGA8A, ITPKA, LTK, DUOXA2, SHF, ONECUT1, ALDH1A2, FOXB1, PIF1, AC069368.3, IGDCC4, SMAD6, SMAD3, SKOR1, CYP11A1, NEIL1, ISL2, CRABP1, KIAA1024, CPEB1, ALPK3, AEN, MESP1, MESP2, LRRK1, MPG, DNASE1L2, KREMEN2, CLDN6, SNN, ERN2, TRIM72, CBLN1, IRX3, IRX5, SLC6A2, GNAO1, MT1E, HERPUD1, GPR56, TEPP, CMTM3, FBXL8, LDHD, NECAB2, C16orf74, IRF8, FOXF1, FOXC2, FOXL1, SLC7A5, ANKRD11, ABR, RTN4RL1, HIC1, TRPV3, VMO1, SLC13A5, TNK1, PER1, HS3ST3A1, HS3ST3B1, PMP22, TEKT3, CCDC144A, RAI1, KCNJ12, SARM1, TMEM132E, MMP28, LHX1, SRCIN1, ARL5C, STAC2, THRA, NR1D1, RAPGEFL1, IGFBP4, PLEKHH3, WNK4, SOST, RUNDC3A, FZD2, GFAP, C1QL1, SP6, HOXB3, HOXB7, PRAC1, HOXB13, CALCOCO2, PHOSPHO1, SPOP, DLX4, SAMD14, ACSF2, USP32, TBX4, LIMD2, CDC42EP4, KIF19, BTBD17, CD300A, OTOP2, EVPL, 9-Sep, TMC6, RP11-1055B8.6, UTS2R, ADCYAP1, TMEM200C, ZNF397, ZNF396, ST8SIA5, SKOR2, SMAD7, LIPG, MBD1, RAX, CDH7, CBLN2, TSHZ1, LINC00908, GALR1, NFATC1, CDC34, PTBP1, SBNO2, MUM1, APC2, ONECUT3, KLF16, GNA11, NFIC, HMG20B, ZNF177, ICAM5, S1PR5, ZNF69, ZNF625-ZNF20, NFIX, LYL1, RFX1, SLC1A6, NOTCH3, BRD4, KLF2, F2RL3, ANO8, SSBP4, ELL, KLHL26, CRTC1, UPF1, ZNF737, CTD-2561J22.3, ZNF43, GRAMD1A, HSPB6, NPHS1, ZNF529, ZNF568, ZNF573, ACTN4, SARS2, L34079.2, LYPD5, CTC-512J12.6, MARK4, VASP, CCDC8, PNMAL1, PPP5D1, ZNF541, GRIN2D, PPFIA3, CTD-3148I10.1, SLC17A7, ZNF577, ZNF615, ZNF614, ZNF880, ZNF808, ZNF701, ZNF83, CACNG8, EPS8L1, ZSCAN5A, ZNF471, ZNF470, AC003005.4, ZNF549, ZIK1, ZNF134, ZNF551, AC003006.7, ZNF814, ZNF418, ZNF256, ZNF606, ZSCAN18, ZNF329, TCF15, RP5-850E9.3, FAM110A, ANGPT4, PROKR2, BTBD3, PCSK2, NKX2-4, NKX2-2, PAX1, FOXA2, CST3, VSX1, DEFB124, XKR7, SOGA1, SLC32A1, TOX2, KCNS1, RBPJL, ZNF334, SALL4, CBLN4, BMP7, ANKRD60, SLC17A9, BHLHE23, UCKL1, ZNF512B, CYYR1, OLIG1, SIM2, TMPRSS2, MICAL3, TBX1, SCARF2, KIAA1671, MN1, TTC28, ZNRF3, RASL10A, NEFH, OSBP2, LGALS2, GALR3, MAFF, FAM83F, MCHR1, WNT7B, CELSR1 |
| Shared by both | PRDM16, TP73, PTCHD2, GRIK3, NTRK1, LHX9, PXDN, DNMT3A, AFF3, FBLN7, DPP10, DLX1, CERKL, SATB2, ALPP, KIAA1257, ZBTB38, KCNAB1, CLDN11, EIF2B5, DGKG, PITX2, SLC6A3, IRX2, CTNND2, UGT3A1, MEF2C, PITX1, ZSCAN12, PNPLA1, TFAP2B, OSTM1, PDE10A, HOXA3, HOXA11, ADCYAP1R1, EPDR1, GRB10, NPTX2, CNPY1, GATA4, CA8, TCF24, NDRG1, BAI1, DMRT1, NPAS4, FLI1, FBRSL1, ZIC2, NKX2-8, NID2, GPR135, CCDC88C, EML1, ZNF598, HS3ST2, TK2, PTRF, TBX21, C18orf42, CELF4, ADAMTS10, PDE4C, ZNF536, RTN2, ZNF667, RIN2, SLC12A5, NFATC2, EDN3 |

**Supplementary table 7. The enriched gene families of NC-DMGs.**

| Gene family | Genes | P value | P adj |
| --- | --- | --- | --- |
| CD molecules | TNFRSF9,TNFRSF8,TNFRSF1B,F11R,TNFSF4,ALK,IL1R1,MME,NCAM1,PLXNC1,SELPLG,FZD10,IL21R,ITGB3,CD300E,TNFSF14,CD79A,C5AR1,CD33,SIGLEC5,LILRB5,LILRB2,LILRA4,LAIR1,LAIR2,LILRA2,LILRB1,KIR3DL1,THBD,ITGB2,IL2RB | 3.50E-05 | 2.76E-02 |
| Zinc fingers C2H2C-type | MYT1L,ST18,MYT1 | 6.61E-04 | 5.22E-01 |
| Receptor tyrosine kinases | NTRK1,ALK,PTK7,MET,ROR2,NTRK3,AATK | 8.18E-04 | 6.45E-01 |
| Calcium voltage-gated channel alpha1 subunits | CACNA1C,CACNA1H,CACNA1A,CACNA1I | 8.98E-04 | 7.09E-01 |
| 5-hydroxytryptamine receptors, G protein-coupled | HTR4 | 3.74E-01 | 1.00E+00 |
| A-kinase anchoring proteins | AKAP6,CBFA2T3 | 2.97E-01 | 1.00E+00 |
| AAA ATPases | PSMC1 | 8.36E-01 | 1.00E+00 |
| Acid sensing ion channel subunits | ASIC2 | 1.82E-01 | 1.00E+00 |
| Activating leukocyte immunoglobulin like receptors | LILRA4,LILRA2 | 2.66E-02 | 1.00E+00 |
| Acyl-CoA synthetase family | SLC27A1,SLC27A5 | 2.35E-01 | 1.00E+00 |

**Supplementary table 8. The enriched gene families of ESCC-DMGs.**

| Gene family | Genes | P value | P adj |
| --- | --- | --- | --- |
| NKL subclass homeoboxes and pseudogenes | HLX,VAX2,EMX1,NOTO,LBX2,TLX2,EN1,DLX1,DLX2,MSX1,HMX1,NKX3-2,NKX6-1,TLX3,NKX2-5, DLX5,EN2,NKX2-6,BARX1,BARHL1,NKX2-3, TLX1,LBX1,VAX1,EMX2,HMX3,HMX2,BSX,NKX2-8,DLX4,NKX2-4,NKX2-2 | 3.27E-21 | 3.06E-18 |
| HOXL subclass homeoboxes | EVX2,HOXD13,HOXD12,HOXD10,HOXD3,HOXD1,GBX2,HOXA2,HOXA3,HOXA9,HOXA11,HOXA13,EVX1,HOXC13,HOXC12,HOXC11,HOXC10,HOXC4,GSX1,PDX1,CDX2,HOXB3,HOXB7,HOXB13 | 5.48E-16 | 5.12E-13 |
| Zinc fingers C2H2-type | PRDM16,GFI1,ZNF648,OSR1,EGR4,GLI2,FEZF2,KLF15,ZBTB38,ZIC4,ZIC1,RREB1,ZSCAN31,ZSCAN12,SP8,ZNF273,FEZF1,ZNF775,ZNF395,OSR2,TRPS1,SCRT1,KLF9,KLF6,ZNF239,WT1,ZIC2,INSM2,BCL11B,KLF13,ZNF598,HIC1,SP6,ZNF397,ZNF396,TSHZ1,KLF16,ZNF177,ZNF69,KLF2,ZNF737,ZNF43,ZNF536,ZNF529,ZNF568,ZNF573,ZNF541,ZNF577,ZNF615,ZNF614,ZNF880,ZNF808,ZNF701,ZNF83,ZSCAN5A,ZNF667,ZNF471,ZNF470,ZNF549,ZIK1,ZNF134,ZNF551,ZNF814,ZNF418,  ZNF256,ZNF606,ZSCAN18,ZNF329,ZNF334,SALL4 | 1.18E-10 | 1.11E-07 |
| Forkhead boxes | FOXE3,FOXD2,FOXL2,FOXF2,FOXP4,FOXH1,FOXD4,FOXB2,FOXN4, FOXG1,FOXA1,FOXB1,FOXF1,FOXC2,FOXL1,FOXA2 | 5.25E-10 | 4.90E-07 |
| Basic helix-loop-helix proteins | HAND2,NEUROG1,HAND1,TFEB,TCF21,OLIG3,TWIST1,TCF24,NEUROG3,MYOD1,NPAS4,MYF6,MYF5,ASCL1,ASCL4,MESP1,MESP2,LYL1,TCF15,BHLHE23,OLIG1,SIM2 | 7.56E-09 | 7.06E-06 |
| LIM class homeoboxes | LHX4,LHX9,LHX6,LHX2,LMX1B,LHX3,LHX5,ISL2,LHX1 | 3.67E-08 | 3.43E-05 |
| PRD class homeoboxes and pseudogenes | PAX7,OTX1,PAX8,PAX3,SHOX2,PHOX2B,PITX2,OTP,PITX1,PAX6,ALX4,ALX1,OTX2,VSX2,GSC,RAX,VSX1 | 9.95E-07 | 9.30E-04 |
| Paired boxes | PAX7,PAX8,PAX3,PAX6,PAX9,PAX1 | 1.24E-05 | 1.15E-02 |
| Endogenous ligands | RSPO1,RGS2,WNT3A,UCN,CCK,TRH,SST,CXCL5,CXCL2,FGF5,RSPO3,NPY,TAC1,RARRES2,PENK,CRH,GDF10,FGF8,CALCB,VWF,WNT1,ADCYAP1,BMP7,EDN3,WNT7B | 2.04E-05 | 1.90E-02 |
| Kruppel like factors | KLF15,KLF9,KLF6,KLF13,KLF16,KLF2 | 2.47E-04 | 2.30E-01 |

**Supplementary table 9. The methylation values of HOXL subclass homeobox genes in normal and pan-cancer samples.**

| Symbol | Mean (Normal) | Sd (Normal) | Mean (Cancer) | Sd (Cancer) | Delta beta |
| --- | --- | --- | --- | --- | --- |
| HOXD10 | 0.2297 | 0.2084 | 0.6031 | 0.1429 | 0.3734 |
| HOXD12 | 0.2227 | 0.1739 | 0.5127 | 0.1029 | 0.2900 |
| HOXD1 | 0.3095 | 0.2061 | 0.5824 | 0.1439 | 0.2730 |
| GSX1 | 0.2787 | 0.0946 | 0.5384 | 0.0931 | 0.2597 |
| HOXC5 | 0.2352 | 0.2203 | 0.4948 | 0.1679 | 0.2596 |
| HOXD8 | 0.3018 | 0.2290 | 0.5599 | 0.1374 | 0.2581 |
| HOXB13 | 0.2173 | 0.0774 | 0.4552 | 0.1384 | 0.2379 |
| PDX1 | 0.2556 | 0.0972 | 0.4619 | 0.0796 | 0.2064 |
| GSX2 | 0.1098 | 0.0729 | 0.2996 | 0.1327 | 0.1899 |
| HOXA2 | 0.5362 | 0.1587 | 0.7165 | 0.1043 | 0.1803 |
| EVX2 | 0.1485 | 0.0663 | 0.3265 | 0.0864 | 0.1780 |
| HOXC12 | 0.4846 | 0.0959 | 0.6550 | 0.0812 | 0.1704 |
| HOXA9 | 0.2845 | 0.0700 | 0.4544 | 0.1143 | 0.1700 |
| HOXB4 | 0.3583 | 0.2103 | 0.5271 | 0.1356 | 0.1688 |
| HOXC13 | 0.2781 | 0.1445 | 0.4414 | 0.1246 | 0.1633 |
| MEOX2 | 0.0902 | 0.0738 | 0.2488 | 0.1154 | 0.1586 |
| HOXD3 | 0.3805 | 0.1272 | 0.5249 | 0.0692 | 0.1444 |
| HOXD4 | 0.4485 | 0.3301 | 0.5916 | 0.1817 | 0.1430 |
| HOXB8 | 0.2657 | 0.1106 | 0.4014 | 0.1407 | 0.1357 |
| HOXC9 | 0.0943 | 0.0372 | 0.2290 | 0.1425 | 0.1347 |
| HOXA5 | 0.6241 | 0.1565 | 0.7564 | 0.0668 | 0.1323 |
| HOXA6 | 0.2145 | 0.1304 | 0.3395 | 0.0824 | 0.1250 |
| HOXD13 | 0.1656 | 0.0625 | 0.2814 | 0.1287 | 0.1158 |
| HOXA7 | 0.2943 | 0.1346 | 0.4084 | 0.1251 | 0.1141 |
| HOXD11 | 0.1804 | 0.0715 | 0.2918 | 0.1037 | 0.1114 |
| HOXC8 | 0.0873 | 0.0669 | 0.1986 | 0.1124 | 0.1113 |
| EVX1 | 0.1356 | 0.2286 | 0.2420 | 0.1918 | 0.1064 |
| MEOX1 | 0.0914 | 0.0636 | 0.1967 | 0.0646 | 0.1053 |
| HOXA4 | 0.4112 | 0.1031 | 0.5072 | 0.1422 | 0.0959 |
| HOXC11 | 0.1610 | 0.1200 | 0.2562 | 0.1410 | 0.0953 |
| HOXD9 | 0.2444 | 0.1815 | 0.3310 | 0.1279 | 0.0866 |
| GBX2 | 0.1980 | 0.0794 | 0.2786 | 0.0798 | 0.0806 |
| HOXA1 | 0.0508 | 0.0360 | 0.1199 | 0.0811 | 0.0690 |
| HOXB6 | 0.7254 | 0.1541 | 0.7826 | 0.0802 | 0.0572 |
| HOXB2 | 0.7629 | 0.1319 | 0.8182 | 0.0869 | 0.0553 |
| HOXB1 | 0.8146 | 0.1758 | 0.8644 | 0.1023 | 0.0498 |
| GBX1 | 0.0470 | 0.0246 | 0.0805 | 0.0390 | 0.0335 |
| HOXA11 | 0.2798 | 0.0654 | 0.3119 | 0.0869 | 0.0321 |
| HOXC4 | 0.4709 | 0.1732 | 0.5015 | 0.1340 | 0.0305 |
| HOXA13 | 0.3964 | 0.1698 | 0.4240 | 0.1930 | 0.0277 |
| HOXC10 | 0.1296 | 0.0682 | 0.1539 | 0.0721 | 0.0243 |
| HOXB9 | 0.1165 | 0.0448 | 0.1289 | 0.0348 | 0.0124 |
| HOXA10 | 0.4462 | 0.1232 | 0.4399 | 0.1250 | -0.0063 |
| MNX1 | 0.1174 | 0.0589 | 0.1086 | 0.0266 | -0.0089 |
| HOXC6 | 0.1052 | 0.0486 | 0.0956 | 0.0327 | -0.0096 |
| CDX4 | 0.6191 | 0.0918 | 0.6047 | 0.0905 | -0.0144 |
| HOXB5 | 0.8190 | 0.1165 | 0.7942 | 0.0672 | -0.0249 |
| CDX1 | 0.6106 | 0.1321 | 0.5738 | 0.1384 | -0.0369 |

**Supplementary table 10. The frequency of methylation events between normal and ESCC samples for CpG sites of *HOXD1* and *GSX1*.** Methylated and unmethylated CpGs were represented by Um and M, respectively.

|  |  | Normal (n) | | ESCC-tissue (n) | | ESCC-cfDNA (n) | |  |
| --- | --- | --- | --- | --- | --- | --- | --- | --- |
|  | CpG site | Um | M | Um | M | Um | M | P value |
| HOXD1 | CpG1 | 6 | 0 | 0 | 18 | 0 | 8 | 1.10E-06 |
|  | CpG2 | 14 | 1 | 0 | 19 | 0 | 12 | 6.25E-11 |
|  | CpG3 | 13 | 3 | 0 | 19 | 3 | 10 | 2.45E-07 |
|  | CpG4 | 16 | 5 | 0 | 19 | 0 | 13 | 2.10E-09 |
|  | CpG5 | 16 | 7 | 0 | 19 | 1 | 12 | 2.21E-07 |
|  | CpG6 | 15 | 8 | 0 | 19 | 0 | 13 | 1.14E-07 |
|  | CpG7 | 16 | 7 | 1 | 18 | 2 | 11 | 1.05E-05 |
|  | CpG8 | 14 | 9 | 0 | 19 | 0 | 13 | 6.23E-07 |
|  | CpG9 | 14 | 9 | 0 | 19 | 0 | 13 | 6.22E-07 |
|  | CpG10 | 14 | 9 | 0 | 19 | 0 | 13 | 6.23E-07 |
| GSX1 | CpG1 | 9 | 14 | 0 | 19 | 2 | 7 | 4.54E-03 |
|  | CpG2 | 10 | 16 | 0 | 19 | 1 | 12 | 1.56E-03 |
|  | CpG3 | 15 | 17 | 0 | 19 | 0 | 13 | 1.76E-05 |
|  | CpG4 | 13 | 20 | 1 | 18 | 0 | 13 | 1.50E-03 |
|  | CpG5 | 11 | 22 | 0 | 19 | 1 | 12 | 3.72E-03 |
|  | CpG6 | 16 | 17 | 0 | 19 | 1 | 12 | 6.44E-05 |
|  | CpG7 | 22 | 11 | 0 | 19 | 0 | 13 | 6.51E-09 |
|  | CpG8 | 18 | 15 | 0 | 19 | 0 | 13 | 1.15E-06 |
|  | CpG9 | 12 | 21 | 0 | 19 | 0 | 13 | 3.69E-04 |
